# Infections and suicide and self-harm: a population-based matched cohort study

**DOI:** 10.64898/2026.06.15.26355702

**Authors:** Jaime Fanjul Iglesias, Georgia R. Gore-Langton, Sharon L. Cadogan, Kathryn E. Mansfield, Ian J. Douglas, Seena Fazel, John Tazare, Caroline Morton, Naaheed Mukadam, Charlotte Warren-Gash

## Abstract

**Background:** Infections have been associated with adverse mental health outcomes, including suicide, but evidence beyond severe or central nervous system infections is limited. We investigated associations between a range of acute infections and subsequent suicide/self-harm outcomes.

**Methods:** We conducted six infection-specific matched cohort studies using English primary care records from the Clinical Practice Research Datalink Aurum (2007-2024), linked to hospital admissions and mortality data. Adults (≥18 years) with a primary care record of infection (gastroenteritis, lower respiratory tract [LRTI], skin/soft-tissue [SSTI], urinary tract [UTI], sepsis, meningitis/encephalitis [positive control]) were matched (age, sex, practice, calendar period) to up to five comparators without infection. We estimated hazard ratios (HRs) for suicide/self-harm outcomes using Cox regression, stratified by matched set and implicitly adjusting for matching factors, with additional adjustment for deprivation, lifestyle factors, and comorbidities. We examined whether associations varied over time, by infection severity, antimicrobial treatment, sex, and prior mental health conditions.

**Findings:** Cohorts ranged from 18,192 individuals with meningitis/encephalitis (matched to 90,915 without) to 398,099 with SSTI (matched to 1,743,747). After adjustment, individuals with infection had a higher hazard of suicide/self-harm outcomes than comparators across all cohorts: sepsis (HR 1.79, 95% CI 1.65-1.93), gastroenteritis (1.62, 1.55-1.70), meningitis/encephalitis (1.56, 1.32-1.84), UTI (1.41, 1.33-1.50), SSTI (1.37, 1.31-1.43), and LRTI (1.37, 1.31-1.44). Risk was highest in the year post-infection, attenuating over time, and was higher among severe infections and those without prior mental health conditions.

**Interpretation:** Common acute infections recorded in primary care are associated with increased risk of suicide and self-harm, particularly following severe infections and in the year post-infection. Findings support suicide risk monitoring following acute infection, particularly among individuals without prior mental health conditions, and highlight infection prevention as a potentially modifiable strategy in vulnerable populations.

**Funding:** Wellcome and La Caixa.

## Background

Suicidal outcomes (suicide/self-harm) represent major and growing public health concerns. In the UK, suicide rates in 2023 reached their highest since 1999, with 11.4 deaths per 100,000 population.(1) Self-harm (defined in the UK as self-poisoning or self-injury irrespective of intent) has increased markedly, from 2.4% of adults aged 16-74 years in 2000 to 10.3% in 2024,(1) and is a strong predictor of near-term suicide.(2) Despite established prevention strategies, progress remains limited by poor risk identification, undertreatment, underreporting, and low prioritisation within public health agendas.(3) Identifying modifiable risk factors is therefore a priority to inform new prevention pathways.

Infections may be one such modifiable risk factor. Infections have been associated with a range of incident mental health conditions.(4) including specific pathogens such as *Toxoplasma gondii*,(5) influenza B,(6) and hepatitis C.(7) Central nervous system infections (meningitis/encephalitis) have documented long-term neuropsychiatric sequelae including anxiety, depression, and severe mental illness.(8–10) Recent evidence has also demonstrated increased risks of mental health conditions following common acute infections more broadly,(11,12) and how critical illness (including severe infections like sepsis) may lead to depression and post-traumatic stress.(13–18) Evidence also links infections to suicide outcomes: in a Danish cohort, hospitalisation for infection was associated with an increased risk of subsequent suicide;(19) similarly, central nervous system (CNS) infections have been linked to elevated suicide risk.(20) While evidence of effect modification in this context is limited, findings from studies of severe illness and hospitalisation more broadly suggest that risk of suicide may vary by infection severity, comorbidity, and time since acute presentation (with risk appearing greatest shortly after hospital discharge).(19,21,22)

However, important evidence gaps remain. Existing evidence is predominantly infection-specific and based on severe, hospital-treated infections. Most infections in England are managed in primary care,(23) yet common, short-lived infections that rarely require hospital care remain understudied, with mixed or inconclusive findings.(19,24) Similarly, the pathways through which infection may increase suicide risk (including whether risk varies by time since infection, infection severity, or demographics) are not well characterised.

Our study aimed to investigate the association between a range of infections (gastroenteritis, lower respiratory tract infections [LRTI], skin/soft-tissue infections [SSTI], urinary tract infections [UTI], sepsis, and positive control exposures (meningitis/encephalitis)) and subsequent suicide and self-harm among adults in England. We also examined whether risk varied with time since infection, infection severity, age, sex, frailty, antimicrobial prescription, or prior mental health conditions.

## Research in context

### Evidence before this study

Prior studies have investigated association between infections and suicide/self-harm outcomes, but evidence has largely focused on severe or central nervous system (CNS) infections identified through hospital records. We searched PubMed on 3 June 2026 using the terms (“infection” OR “infectious disease”) AND (“suicide” OR “self-harm” OR “self-injury” OR “suicidal” OR “suicidality”), restricted to English-language research articles published from 2016 onwards, excluding COVID-19-specific studies. Of the 874 articles identified, and after screening for those relevant to our research question, most investigated CNS infections or hospitalised infections, or examined chronic infections such as HIV or hepatitis C; studies examining common, community-managed acute infections such as gastroenteritis, urinary tract infection (UTI), or skin and soft tissue infection (SSTI) were limited, and the few that existed reported mixed or inconclusive findings. No cohort studies using primary care electronic health records were identified that examined the association between a range of common acute infections and subsequent suicide/self-harm outcomes.

### Added value of this study

To our knowledge, this is the largest cohort study to date examining the association between common acute infections recorded in primary care and subsequent suicide/self-harm outcomes, comprising over 8.1 million participants across six infection cohorts in England. Unlike prior work, which has focused on hospital-treated or CNS infections, our study investigated a wide range of infections predominantly managed in primary care, including gastroenteritis, UTI, SSTI, and lower respiratory tract infections (LRTI), and found elevated hazards of suicide and self-harm across all infections studied. The observation of higher hazards with increased infection severity, the attenuation of risk over time, and the consistent findings across multiple sensitivity analyses strengthen the evidence of a causal effect. The finding that risk was elevated even among individuals without prior mental health conditions identifies a group that may not currently be prioritised for mental health follow-up after infection.

### Implications of all the available evidence

Taken together, the available evidence suggests that common infections, including those managed in primary care, are associated with increased suicide and self-harm. Timely treatment of infections, as suggested by the lower hazards observed in individuals treated with antimicrobials, and prevention strategies such as vaccination may help reduce this risk. Clinicians managing these infections should be aware of the elevated mental health risk in the months following infection, particularly for individuals without prior mental health conditions. Future research should investigate the underlying biological and psychosocial mechanisms, examine the cumulative effect of repeated infections, and assess the effectiveness of targeted mental health interventions following acute infection.

## Methods

### Study design

We conducted a matched cohort study (1 April, 2007-3 April, 2024) using data from the Clinical Practice Research Datalink (CPRD) Aurum (Supplementary Figure 1). This study is reported in accordance with the RECORD checklist (Appendix 1). Feedback from people with lived experience of severe mental illness informed the research objectives and study design.

CPRD Aurum is a large, broadly representative database of pseudonymised English primary care records (recorded using SNOMED-CT morbidity coding).(25) Linked Hospital Episode Statistics (HES) Admitted Patient Care records,(26) capture national inpatient hospital admissions. Cause of death was obtained from linked Office for National Statistics (ONS) mortality records. The Index of Multiple Deprivation (IMD) is an area-based composite measure of relative socioeconomic deprivation for small geographic areas in England.(27)

Our study was approved through CPRD’s Research Data Governance process (Ref 24_003746), and locally by the London School of Hygiene and Tropical Medicine Research Ethics Committee (Ref 30490).

### Participants

We included individuals ≥18 years who were registered with a CPRD Aurum general practice for at least 12 months before study entry and from practices contributing research-standard data during the study period. Participants were eligible for inclusion from the latest of: study start (1 April, 2007), one year after registration with their practice, or their 18th birthday.

For each infection, we identified an exposed cohort comprising individuals with a recorded morbidity-coded (using SNOMED-CT coding) diagnosis of that specific infection during the study period. For gastroenteritis, we also accepted a symptom or pathogen code. Individuals could contribute to more than one infection cohort. Each exposed individual was randomly matched (without replacement) to up to five unexposed individuals from the general population on age (within 2 years), sex, and primary care practice. We matched unexposed individuals in chronological order of exposed individuals’ cohort entry dates to minimise time-related biases. We defined index date for exposed individuals as the date of first recorded infection during the study period. Each matched set had to include at least one exposed individual and at least one unexposed individual. Due to very large numbers of individuals with common infections recorded in primary care, we drew a random 10% sample of individuals with SSTI, UTI, gastroenteritis, and LRTI prior to matching; individuals not selected remained eligible as comparators until their infection date, when they were censored.

The index date for comparators was the date of the matched exposed individual’s infection record. In each infection cohort, unexposed participants who subsequently received a diagnosis of the infection after their index date were censored from the unexposed group and became eligible for inclusion in the corresponding exposed cohort, where they were matched with up to five new unexposed comparators.

Participants were followed from cohort entry until the earliest of: first recorded outcome, death (from causes other than suicide), end of practice registration, last practice data collection date, or study end (3 April, 2024). We excluded individuals with <1 day of follow-up.

### Procedures

Code lists for all exposures, outcomes, and covariates are available online.(28) Our primary outcome was a composite of the earliest (since index date) of: a first primary care diagnostic code for self-harm (any act of self-poisoning or self-injury irrespective of intent), or a suicide death identified from ONS mortality records (ICD-10 coded). As a secondary analysis, self-harm and suicide outcomes were examined separately.

We selected covariates *a priori* based on a directed acyclic graph (Supplementary Figure 2). We measured covariates at or before cohort entry except for smoking status and obesity (covariate definitions: Supplementary Table 1). Covariates comprised: age (categorised: 18-29, 30-39, 40-49, 50-64, ≥65 years); sex (female/male); ethnicity (using a previously validated algorithm and categorised as: White, South Asian, Black, Mixed, Other, Missing);(29) deprivation (quintiles of individual-level IMD, supplemented when missing with practice-level data); lifestyle factors (harmful alcohol use, smoking status, obesity); comorbidities (Charlson Comorbidity Index [CCI], common mental disorders [CMD; depression, anxiety], severe mental illness [SMI; bipolar disorder, schizophrenia, other psychoses], pre-index date self-harm); and healthcare utilisation (number of primary care consultations in the year prior to index date). We included recreational injectable drug use as an additional covariate in the SSTI cohort only.(30) Smoking status and obesity status (BMI ≥30 kg/m²) were derived from primary care records using the record closest to the index date, and treated as missing if unrecorded. We assessed frailty using the electronic Frailty Index (eFI), which categorises individuals as fit, mildly frail, moderately frail, or severely frail based on 36 deficits recorded in primary care.(30)

## Statistical analysis

### Main analysis

We described the characteristics of participants in each infection cohort by exposure status and including missing data for each variable. We estimated adjusted rate differences (95% confidence intervals) in outcomes between infected and uninfected participants for each cohort.

We estimated hazard ratios (HRs) for the association between infection and combined suicide/self-harm outcomes using Cox proportional hazards regression stratified by matched set. We calculated follow-up time (underlying timescale in all Cox models) in years, from index date to the earliest of: outcome date, death, end of registration, end of the study period, or, among comparators, record of the cohort-specific infection. We fitted sequential models for each infection cohort, using complete-case analysis: a minimally adjusted model stratified by matched set (implicitly adjusting for age, sex, practice, and calendar period); Model 1, additionally adjusted for deprivation; Model 2, further adjusted for lifestyle factors (including injectable drug use in the SSTI cohort); Model 3, further adjusted for comorbidities; and a fully adjusted model additionally adjusting for ethnicity (as a sensitivity analysis, due to high proportions of missing ethnicity data).

### Sensitivity analyses

We conducted six prespecified sensitivity analyses: 1) restricting of the study period to before 1 March 2020 to investigate the impact of including COVID-19 pandemic time in main analyses; 2) restricting follow-up to begin one year after the index date, to limit outcome ascertainment bias and reverse causation; 3) restricting to participants with at least one primary care consultation in the year before the index date to exclude practice non-attenders; 4) an additionally adjusted model with missing ethnicity retained as an unknown category; 5) restricting to participants with no self-harm episodes in the two years prior to the index date, to assess whether findings were robust to excluding self-harm episodes that may represent continuation of a pre-existing episode rather than a new infection-associated event; and 6) examining self-harm and suicide as separate outcomes to understand their individual contributions to the composite.

### Secondary analyses

To assess whether risk varied over time, we fitted the fully adjusted Cox model on cumulative follow-up windows of 0-1, 0-2, 0-3, 0-4, and 0-5 years after index date, and a likelihood ratio test compared a model with a main exposure effect against one including a time-interaction term.

We assessed whether age, sex, frailty, and prior mental health conditions modified the effect of infections on suicide/self-harm using likelihood ratio tests comparing models with and without interaction terms and estimating stratum-specific hazard ratios. Our frailty analysis was restricted to participants aged 65 years or older, the age group in which the frailty index has been validated.(30) Prior mental health concerns was defined as a composite binary variable (any pre-index record of CMD, SMI, or self-harm).

To examine the role of infection severity, we created a three-level exposure restricting to participants eligible for HES-linkage: without infection, ‘non-severe’ infection (CPRD record only), or ‘severe’ infection (CPRD record plus HES record of hospital admission of the same infection or sepsis 28 days either side of the CPRD record). We also investigated antimicrobial prescribing within 7 days of index date, creating a three-level exposure of unexposed, untreated infection, and treated infection. We conducted both analyses in the gastroenteritis, LRTI, SSTI, and UTI cohorts only, as meningitis/encephalitis and sepsis were assumed severe in all cases and hospital-administered antimicrobials are not captured in CPRD.

We conducted all analyses in R (version 4.4.1).

## Results

We included: 311,917 individuals with gastroenteritis matched to 1,405,871 without; 379,975 with LRTI matched to 1,627,978 without; 398,099 with SSTI matched to 1,743,747 without; 192,520 with UTI matched to 874,703 without; 190,913 with sepsis matched to 947,951 without; and 18,192 with meningitis/encephalitis matched to 90,915 without (Table 1, Supplementary Figures 3a/3b). Median follow-up ranged from 2.0 years (interquartile range [IQR]: 0.5-4.8) in the sepsis cohort to 6.0 years (IQR: 2.3-10.7) in the LRTI cohort (Table 1). Baseline characteristics are shown in Table 1. Compared to uninfected comparators, individuals with infection had higher rates of suicide/self-harm (Supplementary Figure 4), comorbidity, prior mental health conditions, and primary care consultations in the year before cohort entry across all cohorts. Compared to those with missing covariate data, individuals included in complete-case analyses were younger and more likely to be male across all cohorts (Supplementary Table 2).

**Table 1.**
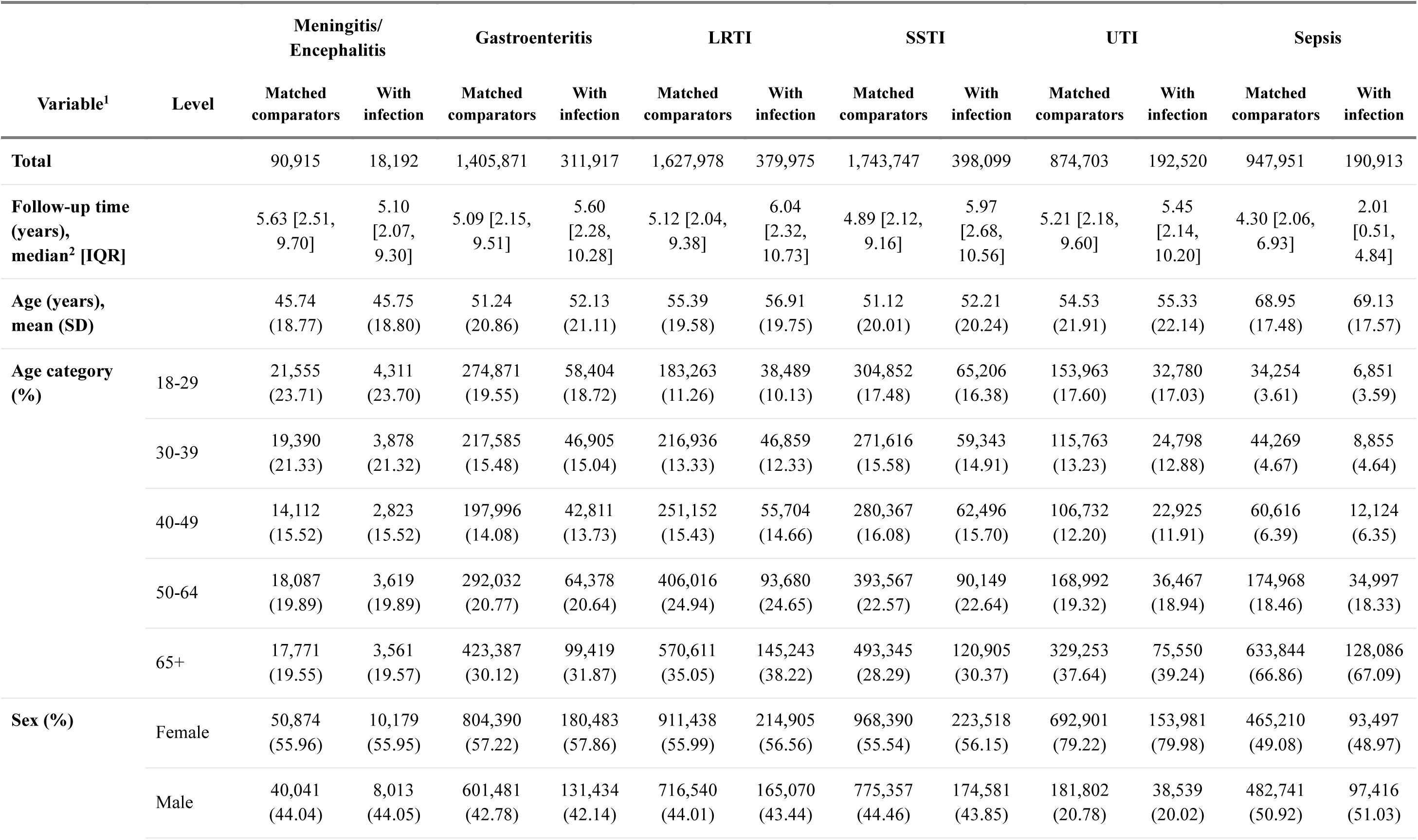
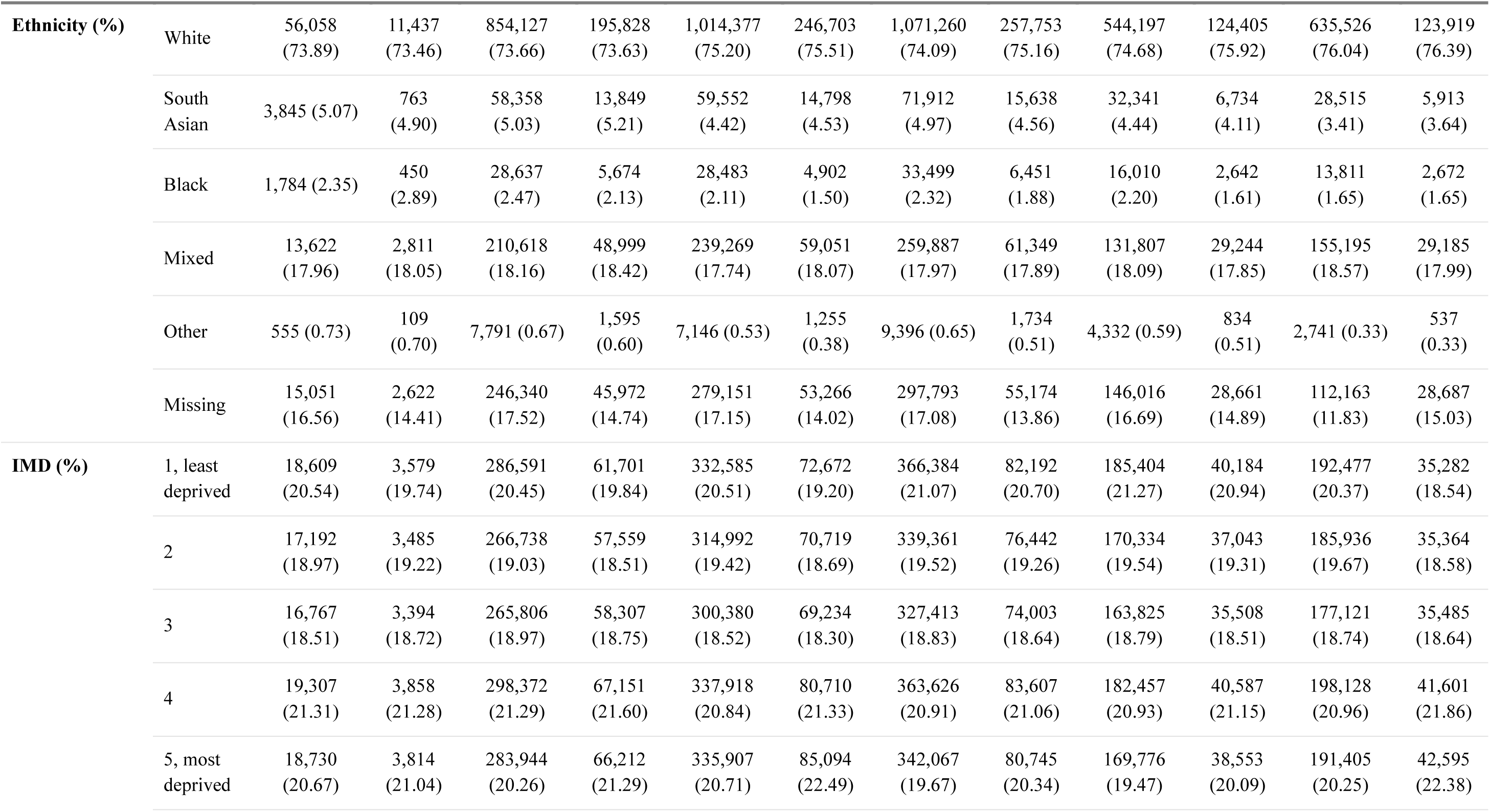
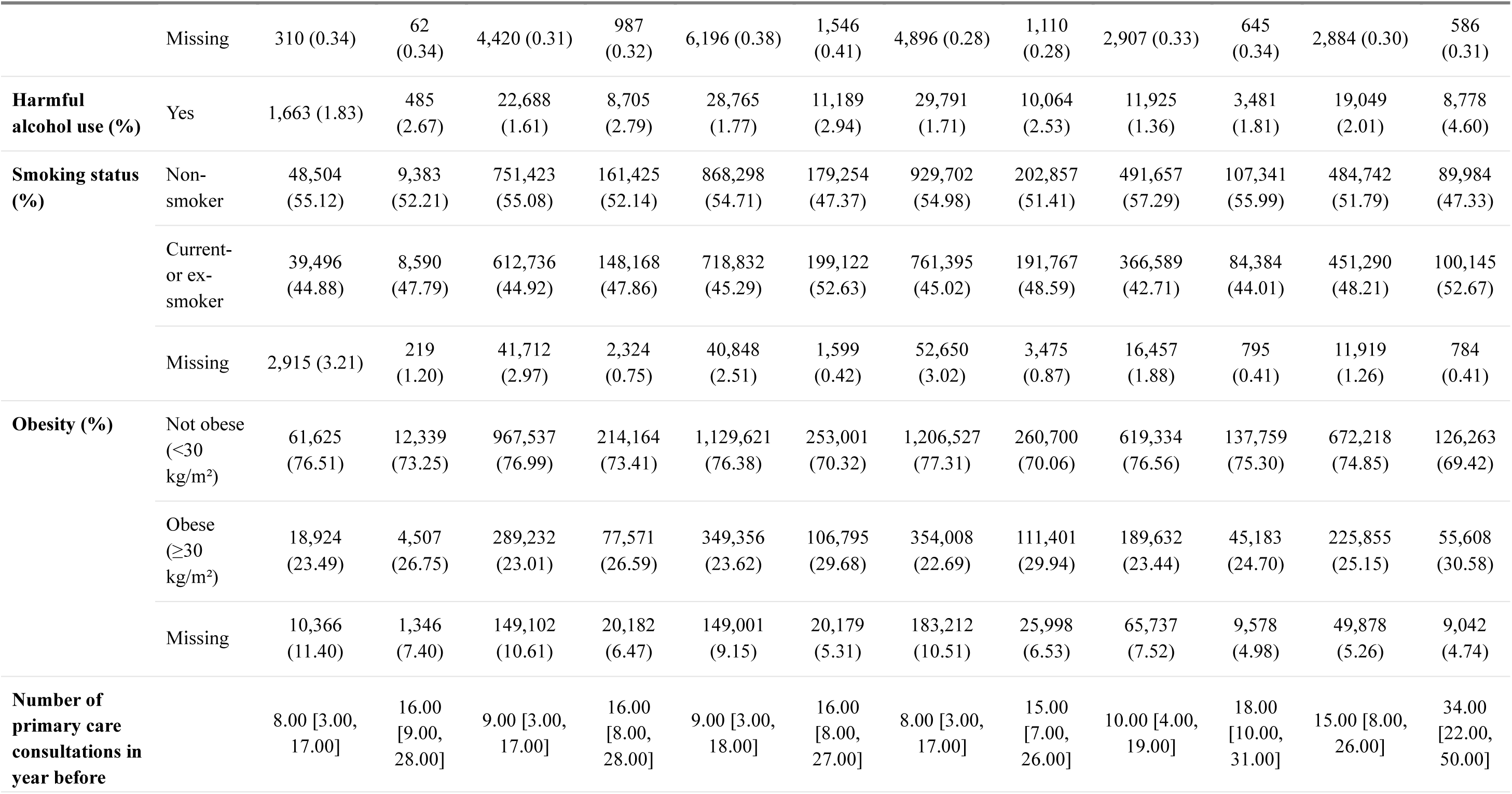
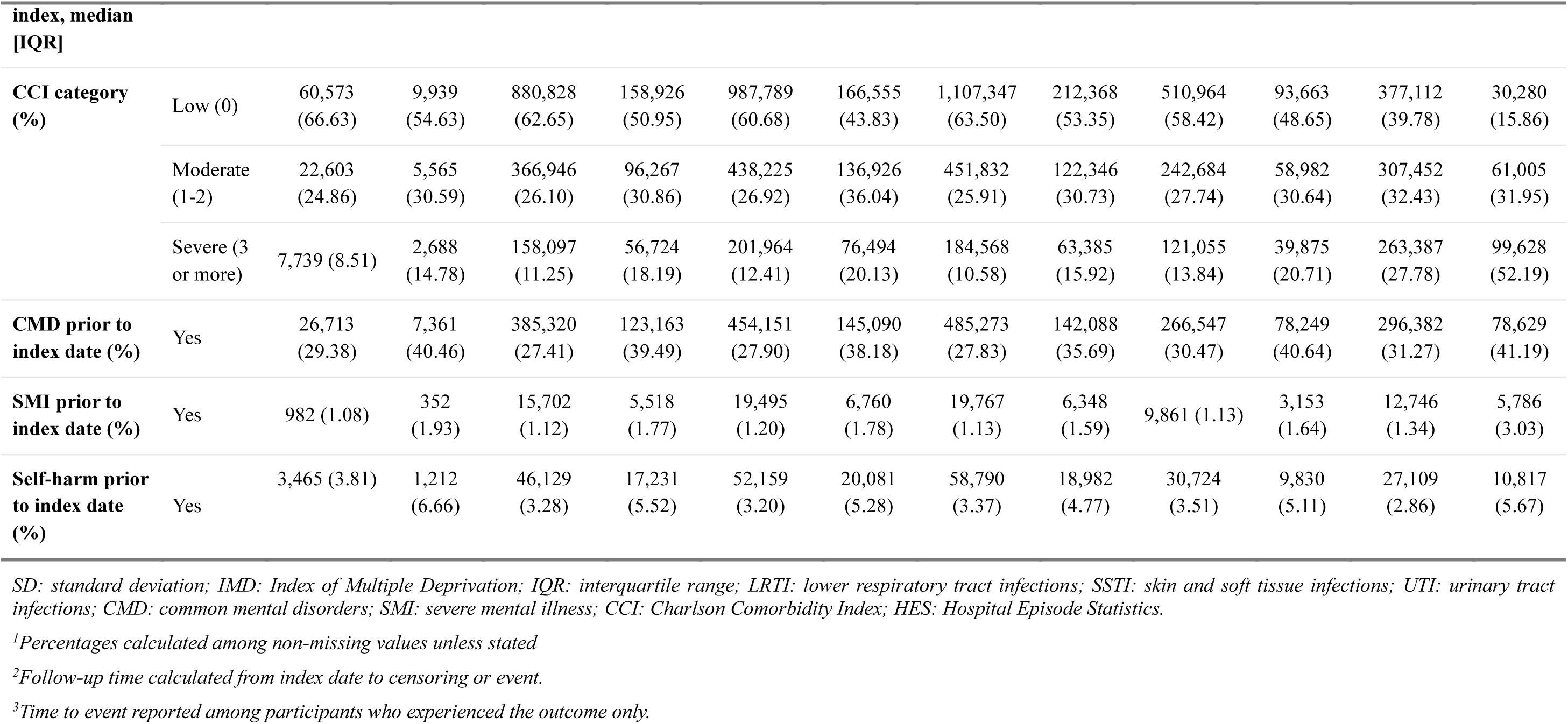
Baseline characteristics and follow-up summary of participants by infection status (figures are n [%] unless otherwise stated)

Rate differences in suicide/self-harm outcomes between infected and uninfected participants ranged from 0.54 (95% CI 0.47-0.62) per 1,000 person-years in the LRTI cohort to 1.43 (1.28-1.59) in the sepsis cohort (Supplementary Table 3). Kaplan-Meier curves showed higher cumulative incidence of suicidal outcomes in infected participants across all cohorts from early in follow-up (Supplementary Figure 5).

In fully adjusted Cox models, participants with infection had higher hazards of suicide/self-harm outcomes than uninfected comparators across all six cohorts (Figure 1). Associations were strongest for sepsis (HR 1.79, 95% CI 1.65-1.93) and gastroenteritis (1.62, 1.55-1.70), followed by meningitis/encephalitis (1.56, 1.32-1.84), UTI (1.41, 1.33-1.50), SSTI (1.37, 1.31-1.43), and LRTI (1.37, 1.31-1.44). Attenuation from minimally to fully adjusted estimates was driven largely by adjustment for comorbidities and prior mental health conditions and was strongest for sepsis and gastroenteritis (Supplementary Figure 6). Point estimates were consistent in the model additionally adjusted for ethnicity, and across all sensitivity analyses (Supplementary Table 4). When outcomes were examined separately, self-harm associations closely mirrored the composite results across all cohorts; for suicide, evidence of an association was observed only for UTI (HR 1.83, 95% CI 1.22-2.74) and sepsis (1.84, 1.20-2.80), with imprecise estimates for the remaining cohorts (Figure 1, Supplementary Table 4).

**Figure 1.**
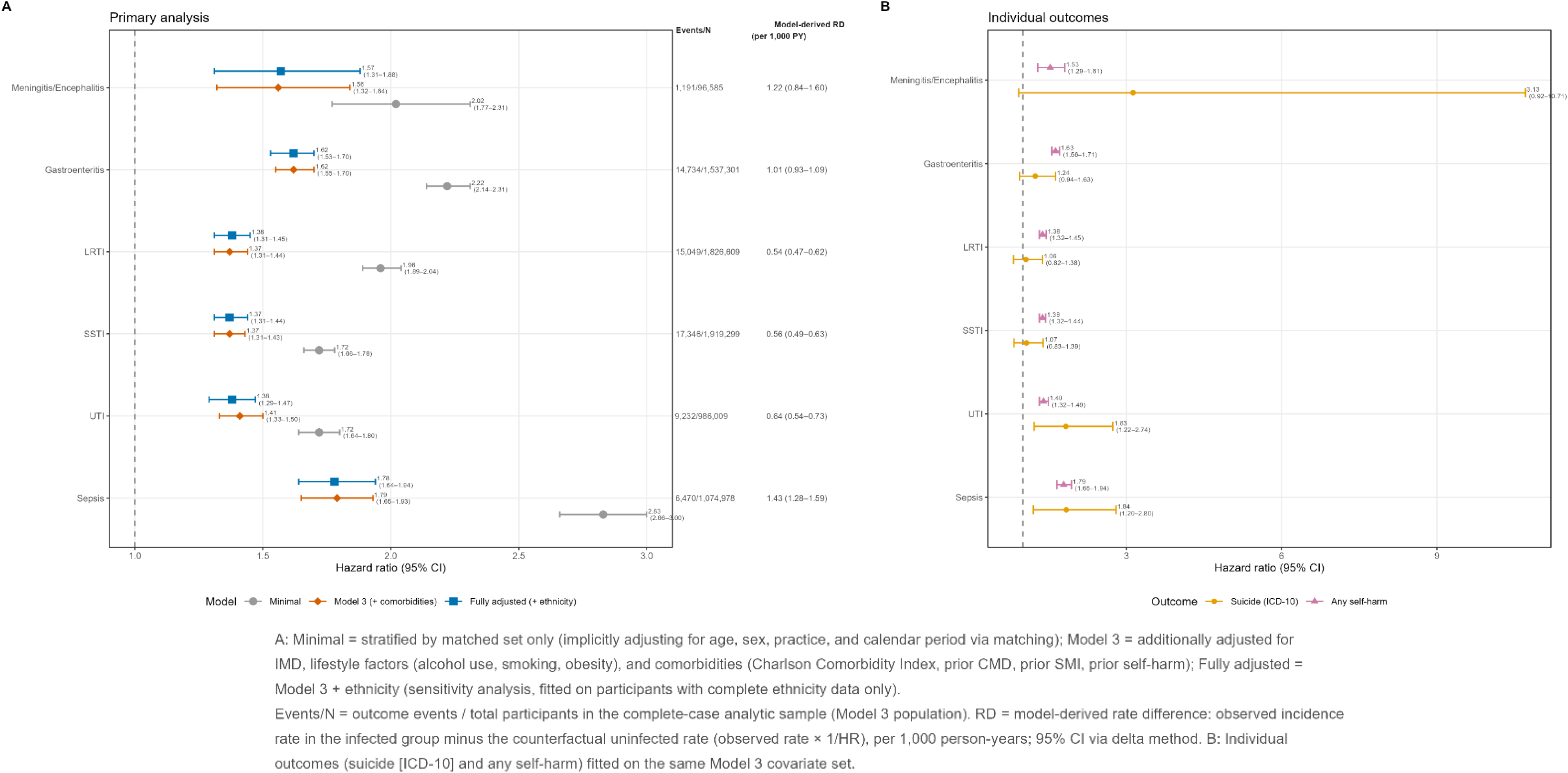
Forest plot of minimally adjusted, fully adjusted and additionally adjusted hazard ratios for each infection (composite and individual outcomes)

There was strong evidence that the association varied over follow-up time in all six cohorts (p<0.0001 for all). Hazard ratios were highest in the first year post-infection and attenuated but persisted up to five years in most cohorts, with the exception of LRTI where attenuation was less evident (Figure 2, Supplementary Table 5). Rate differences in the first year post-infection ranged from 0.95 (95% CI 0.69-1.22) per 1,000 person-years in the LRTI cohort to 3.78 (2.47-5.09) in the meningitis/encephalitis cohort (Supplementary Table 6).

**Figure 2.**
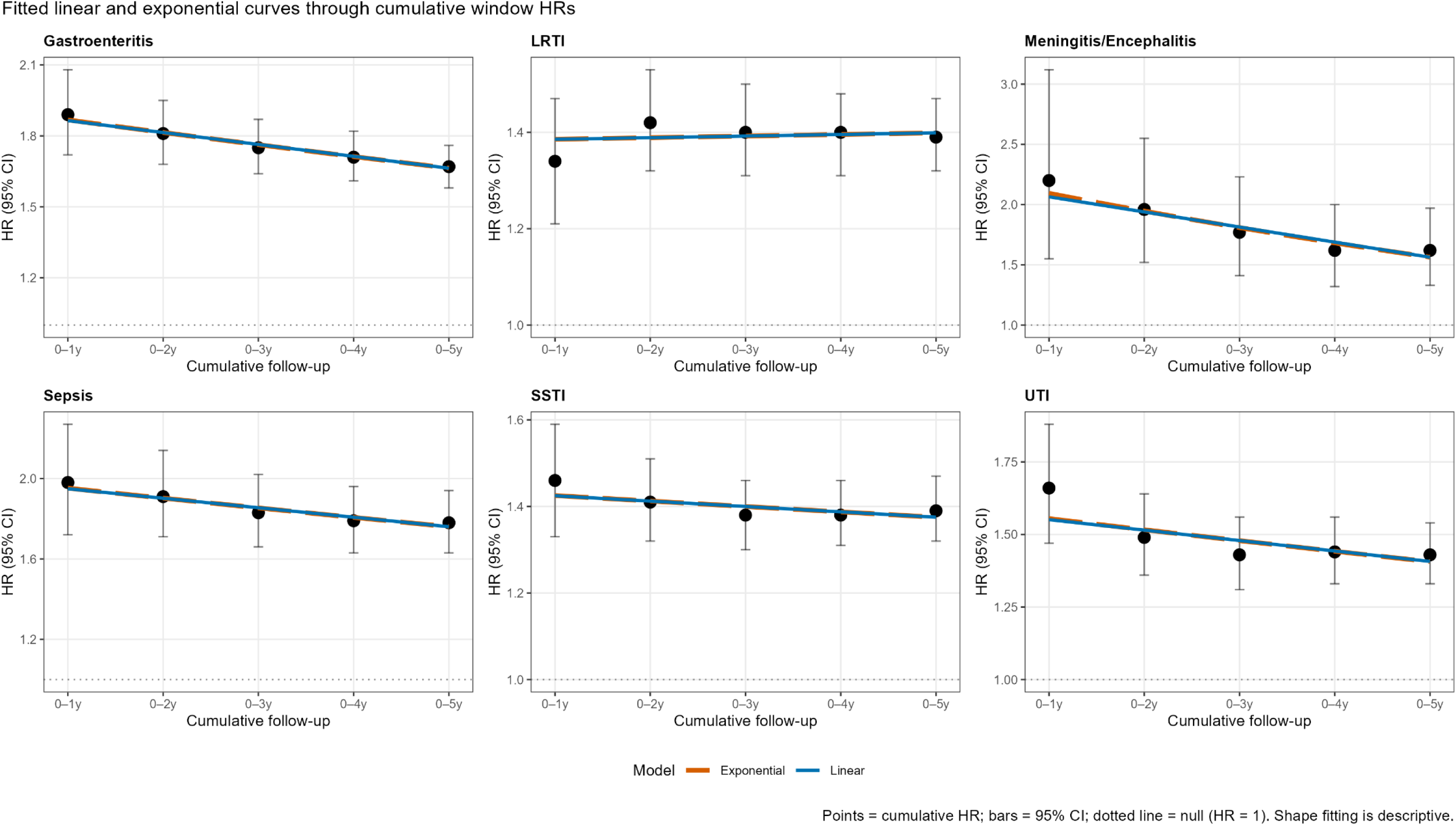
Adjusted yearly estimates of the hazard ratio for infection on suicide/self-harm, estimated by Cox regression.

There was strong evidence that prior mental health conditions modified the association across all six cohorts (p<0.0001 for all except meningitis/encephalitis, p=0.031), with higher hazards among those without prior mental health conditions; the difference was most pronounced for sepsis (HR 2.05 [1.79-2.35] without vs 1.66 [1.51-1.83] with) and meningitis/encephalitis (1.84 [1.35-2.50] vs 1.43 [1.16-1.76]) (Table 2). There was evidence of effect modification by sex only in the UTI cohort (p=0.023), where men showed a stronger association than women (1.69 [1.43-1.99] vs 1.37 [1.29-1.47]). There was statistical evidence of frailty modification in most cohorts (p<0.0001) but stratum-specific estimates were inconsistent in direction and imprecise, particularly for meningitis/encephalitis (p=0.32). There was little evidence of effect modification by age in any cohort.

**Table 2.**
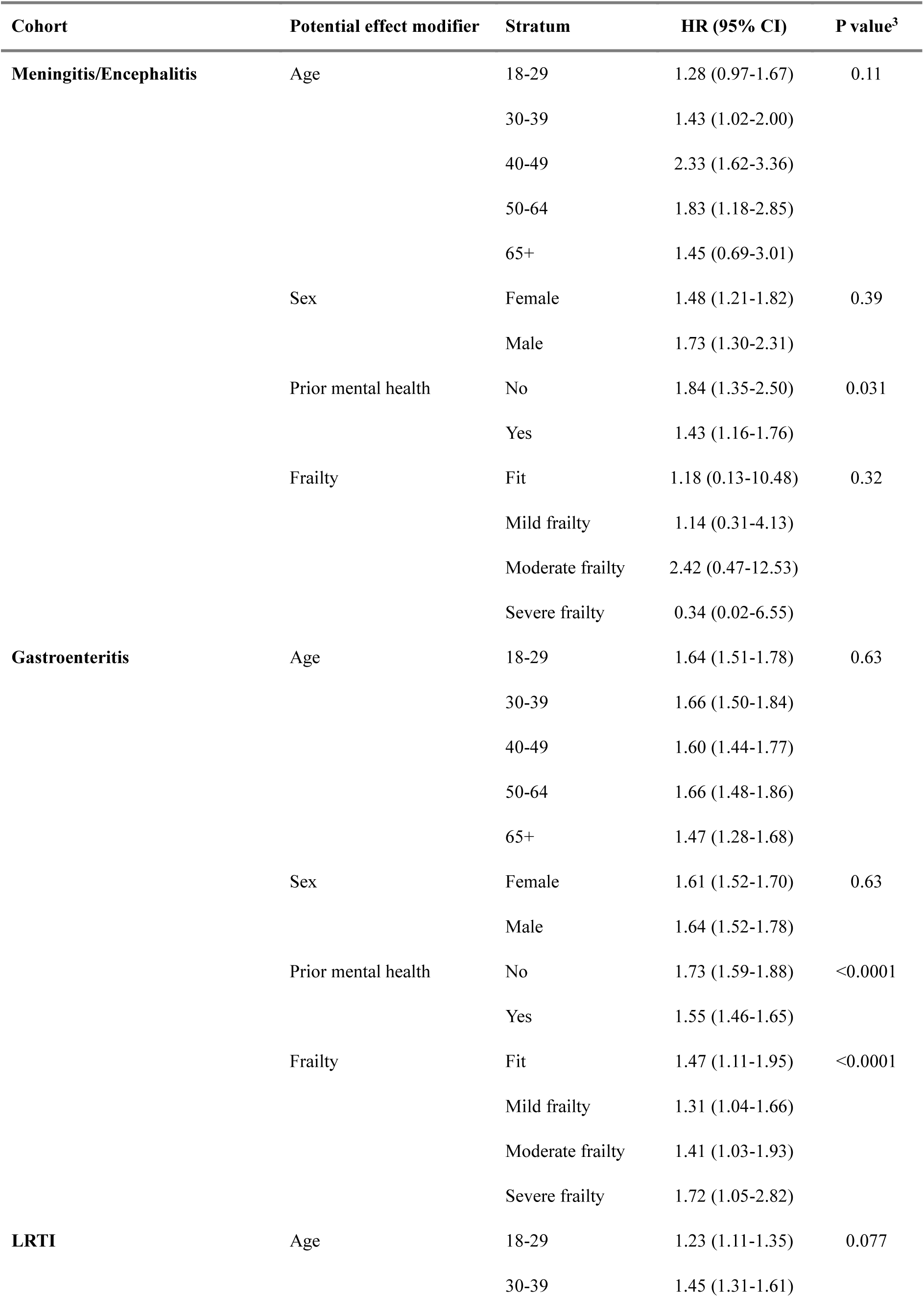
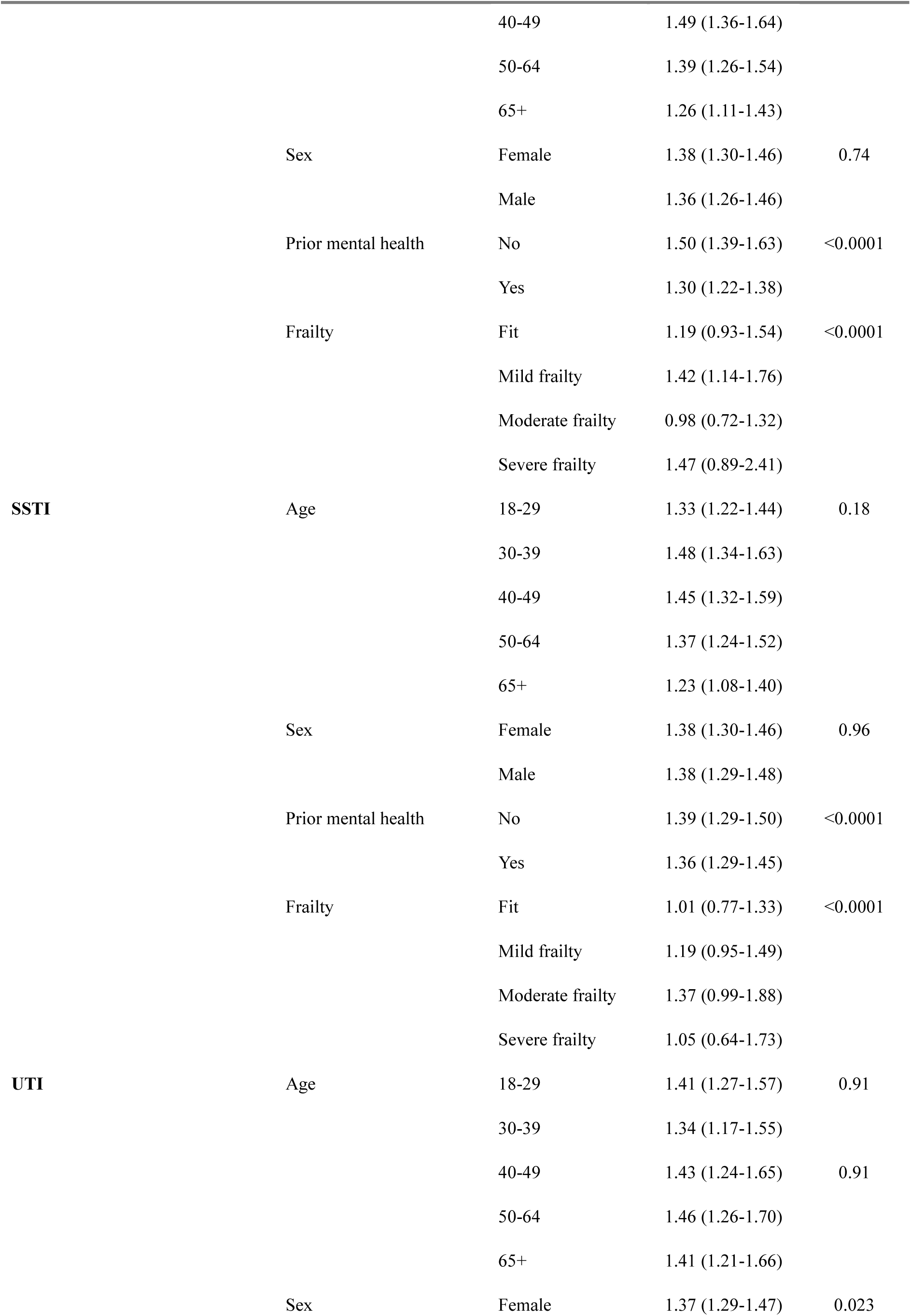
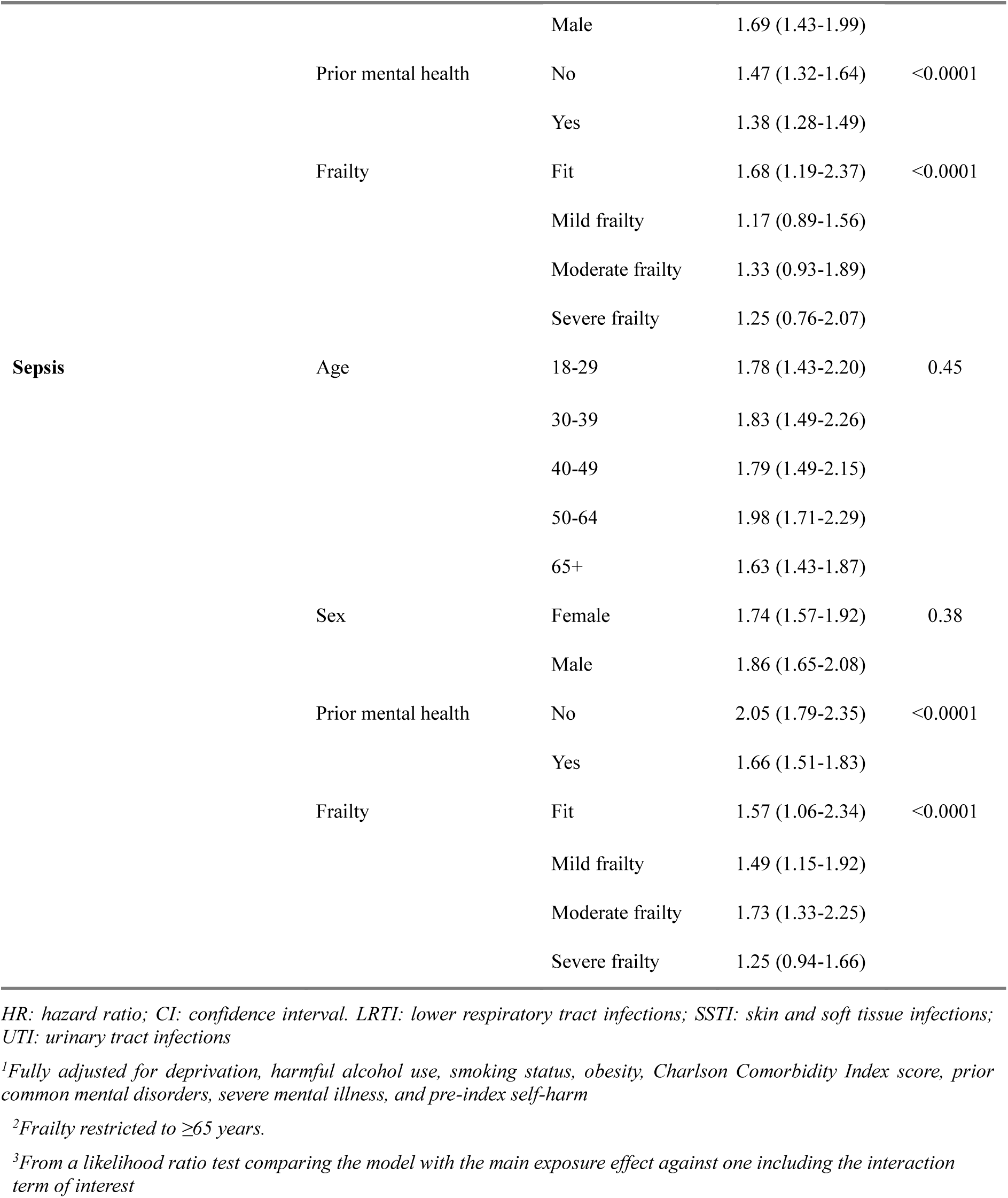
Adjusted^1^ hazard ratios (95%CI) for the association between infection and suicide/self-harm by age, sex, frailty^2^, and prior mental health.

There was evidence of a severity gradient across all four cohorts, most clearly for LRTI (HR 1.34 [1.26-1.41] non-severe vs 2.19 [1.75-2.75] severe); a similar directional pattern was observed for UTI, SSTI, and gastroenteritis, though confidence intervals overlapped (Figure 3).

**Figure 3.**
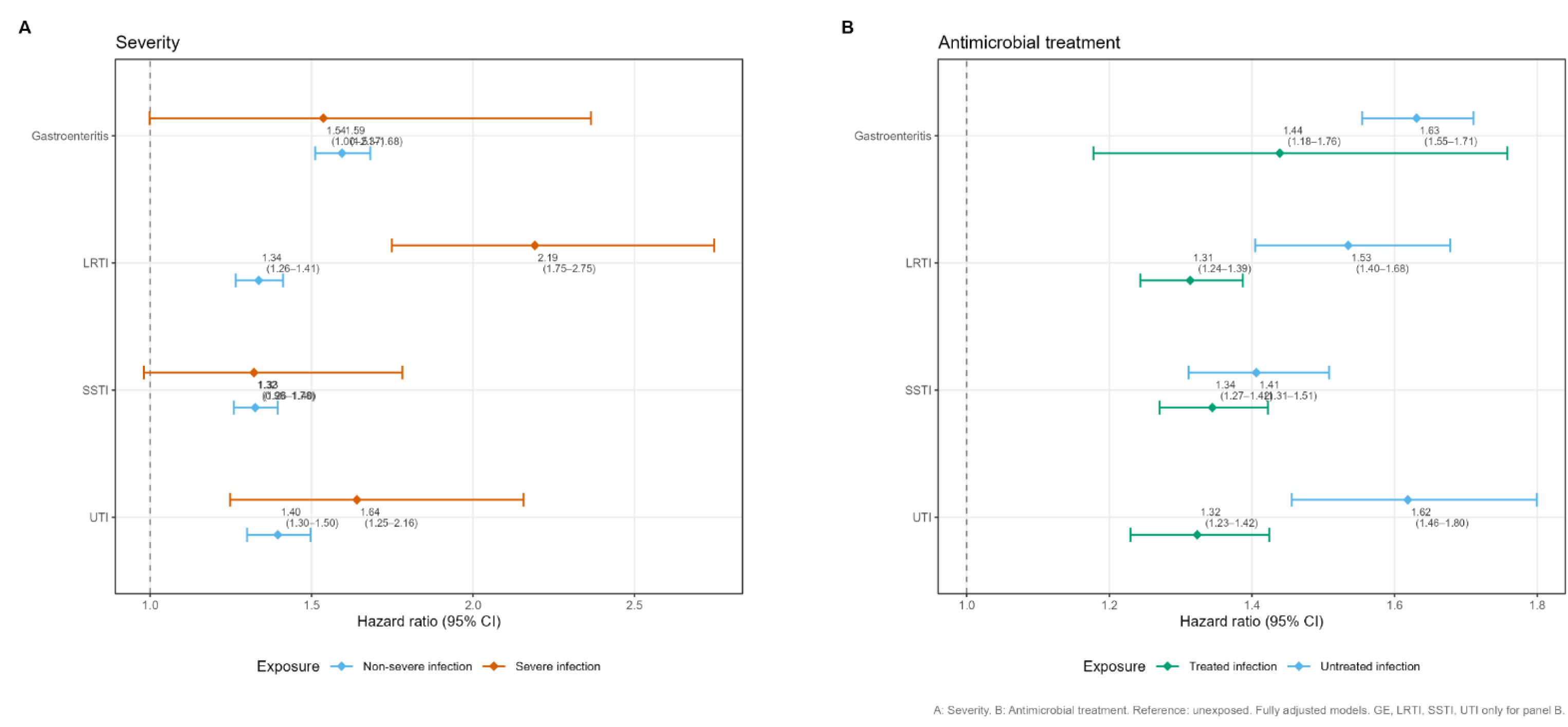
Forest plot of infection severity and antimicrobial treatment analysis (adjusted HRs)

Antimicrobial treatment was associated with lower hazards than untreated infection across all four cohorts, most clearly for LRTI (1.53 [1.40-1.68] untreated vs 1.31 [1.24-1.39] treated) and UTI (1.62 [1.46-1.80] vs 1.32 [1.23-1.42]); hazard ratios remained elevated above 1.0 in both groups (Figure 3).

## Discussion

For over 8 million individuals across six infection cohorts followed for up to 17 years, adults with a primary care record of an acute infection had higher hazards of suicide/self-harm outcomes than uninfected comparators, with rate differences in the year post-infection ranging from 0.95 to 3.78 per 1,000 person-years. Risk was highest in the first year, attenuating over time, and was stronger for more severe infections, among those without prior mental health conditions, and in men with UTI; lower hazards were observed in treated versus untreated infections, though this was not consistent across all infection cohorts.

The finding of elevated risk for meningitis/encephalitis, used here as a positive control, is consistent with prior Danish registry studies reporting increased suicide risk following CNS infections.(19,20) For the other infections, the direction of association was also consistent with the broader prior evidence linking hospitalised infections to subsequent suicide risk,(19,22) though effect estimates in our study are generally lower, likely because prior registry studies captured hospital-treated infections representing more severe infections, while our exposure included specific primary care-recorded infections, consequently representing a broader group of individuals and including those less likely to have severe infection.

Higher hazards with greater infection severity, most clearly for LRTI, support two explanatory mechanisms. First, more severe infections are associated with greater systemic inflammation and higher cytokine responses; inflammatory cytokines can influence mood regulation and suicidality through effects on neurotransmitter systems.(31) Second, severe infections are associated with psychosocial stressors including perceived life threat, prolonged recovery, and chronic sequelae, all of which may independently increase suicide and self-harm risk.(22) The attenuation of risk over time is consistent with both mechanisms, as acute inflammatory responses and illness-related stressors are most intense immediately following infection.

Unlike the other cohorts, non-severe gastroenteritis was associated with higher hazards than severe gastroenteritis, though the severe estimate was imprecise (HR 1.54, 95% CI 1.00-2.37). This likely reflects that most mild gastroenteritis may not prompt a healthcare consultation,(32) meaning primary care-recorded cases (even those without a hospital admission) may already represent a relatively severe subset. Additionally, gut-brain axis pathways, including microbiota perturbation following enteric infection, have been proposed as a mechanism linking gastrointestinal infection to mental health outcomes,(32) which could contribute to elevated risk even in the absence of severe systemic illness.

Lower hazards among infections with a record of antimicrobial treatment could reflect a biological effect of earlier treatment reducing the severity and duration of the inflammatory response, greater healthcare engagement facilitating earlier identification of mental health needs, or the inclusion of hospital-treated severe infections in the untreated group (for which primary care prescribing data are unavailable in CPRD). That hazards remained above 1.0 in both treated and untreated groups suggests the association persists regardless of antimicrobial treatment, and that additional mental health support may be warranted in either case.

There was strong evidence that prior mental health conditions modified the association between infection and suicide/self-harm outcomes, with higher hazards among those without prior mental health conditions. Individuals with prior mental health conditions carry a substantially elevated baseline risk,(33) such that the contribution of an acute infection episode may be comparatively small (though hazards remained elevated in this group too). Rather, infection appears a proportionally greater trigger in those without a pre-existing psychiatric history, and prior mental health status could assist in risk stratification for targeted post-infection support.(34)

The observation that effect modification by sex was seen only in the UTI cohort, where men showed a stronger association than women, warrants cautious interpretation. The composite outcome combines self-harm, more commonly recorded in women, and suicide deaths, more frequently completed by men due to use of more lethal methods rather than higher attempt rates.(1,3) The observed sex difference may therefore reflect differential healthcare-seeking or recording of self-harm rather than a true difference in underlying risk.

Evidence of effect modification by frailty was observed in most cohorts, but stratum-specific estimates were inconsistent in direction across infections and without a clear gradient, limiting interpretation despite prior evidence linking frailty to increased suicidal behaviour in older adults.(35)

Our study used data from a large, nationally representative primary care dataset, providing substantial statistical power and good external validity. Linked cause-specific mortality data ensured capture of suicide deaths not recorded in primary care. The infections investigated varied widely in severity, anatomical site, and clinical course, and the positive control design using meningitis/encephalitis offered an internal validity check against prior literature. Primary care records limited recall bias, and multiple sensitivity analyses yielding consistent results support the robustness of our findings.

There were several limitations. We retained individuals with prior self-harm in the main analysis on the assumption that episodes separated by sufficient time represented independent events rather than continuation of an incident condition; sensitivity analyses restricting to those with no self-harm in the two years prior to index date yielded consistent results. Self-harm is likely substantially under-ascertained in primary care, and this may be differential; individuals consulting their GP with an infection may also be more likely to have self-harm recorded, potentially leading to overestimated results. The composite outcome was predominantly comprised of self-harm episodes, limiting extrapolation specifically to suicide deaths. Primary care-coded infections likely represent more symptomatic presentations, meaning effect estimates may be overestimates relative to the true population-level association; conversely, infections in comparators not recorded in primary care would bias estimates toward the null. Despite adjusting for a wide range of confounders, residual confounding remains possible, particularly from unmeasured social stressors and healthcare-seeking preferences. Time-varying covariates assessed at baseline, such as CCI, may not reflect changes over extended follow-up but avoids adjusting for factors which may lie on the causal pathway. Cumulative effects of repeated infections were not examined. Finally, generalisability to non-UK healthcare settings may be limited in the absence of specific mechanistical evidence.

Future research should examine cumulative effects of repeated infections, use laboratory-confirmed definitions, distinguish suicide from self-harm, and clarify inflammatory and psychosocial mechanisms to strengthen causal interpretation.

Clinicians managing common acute infections should be aware that individuals (particularly those without prior mental health conditions, those without antimicrobial treatment, and men with UTI) may be at increased risk of self-harm and suicide in the months following infection. Mental health monitoring post-infection could help identify at-risk individuals,(36) though whether small absolute rate differences justify routine screening warrants further evaluation. Within mental health services, enquiring about recent infection history may identify at-risk individuals for targeted prevention strategies.

## Contributors

CWG conceptualised the study design, secured funding, curated the data, and administered the project, resources, and software. CWG and GRGL verified the underlying data. JFI led the investigation, with support from CWG and GRGL. JFI conducted the formal analysis, alongside input from CWG and GRGL. Supervision and validation were led by CWG and GRGL. JFI wrote the original manuscript, which was reviewed and edited with input from all authors. CWG is the guarantor of the study. KEM developed the study design and reviewed the manuscript. All authors interpreted the results, had full access to all the data in the study, and had final responsibility for the decision to submit for publication.

## Data sharing

Electronic health records are considered sensitive data in the UK by the Data Protection Act and cannot be shared by public deposition because of information governance restriction in place to protect patient confidentiality. Access to data is available only once approval has been obtained through the individual constituent entities controlling access to the data. Data from the primary care practices can be requested with an application to the Clinical Practice Research Datalink (CPRD; https://www.cprd.com/).

## Acknowledgements

This study is based in part on data from the Clinical Practice Research Datalink obtained under licence from the UK Medicines and Healthcare products Regulatory Agency. The data is provided by patients and collected by the NHS as part of their care and support. The interpretation and conclusions contained in this study are those of the author/s alone. The study was approved by the CPRD’s research data governance process (Protocol number: 24_003746).

