## Supplementary Material for "Infections and suicide and self-harm: a population-based matched cohort study"

Appendix 1. RECORD checklist for cohort studies

Supplementary Table 1. Variable definitions

Supplementary Table 2. Missing data summary and complete case comparison

Supplementary Table 3. Model-derived rate differences (per 1,000 person-years) between infected and uninfected participants (full follow-up period)

Supplementary Table 4. Sensitivity analyses results

Supplementary Table 5. Association between infection and suicide/self-harm across cumulative follow-up windows

Supplementary Table 6. Model-derived rate differences (per 1,000 person-years) between infected and uninfected participants (first-year post-infection)

Supplementary Table 7. Additional variable information

Supplementary Figures

Supplementary Figure 1. Study design diagram

Supplementary Figure 2. Directed acyclic graph illustrating hypothesized confounding relationships between infection and suicide or self-harm

Supplementary Figures 3a/3b. Flowcharts illustrating participant identification

Supplementary Figure 4. Suicidal outcome events by infection group

Supplementary Figure 5. Kaplan-Meier plots comparing the survival for suicide/self-harm of patients with and without an infection

Supplementary Figure 6. Model attenuation: hazard ratios for infection on suicide/self-harm across adjustment models

#### Appendix 1. RECORD checklist for cohort studies

| | Item No. | STROBE items | Location in manuscript where items are reported | RECORD items | Location in manuscript where items are reported |
|---|---|---|---|---|---|
| **Title and abstract** | | | | | |
| | 1 | (a) Indicate the study's design with a commonly used term in the title or the abstract (b) Provide in the abstract an informative and balanced summary of what was done and what was found | Title; Abstract | RECORD 1.1: The type of data used should be specified in the title or abstract. When possible, the name of the databases used should be included.<br><br>RECORD 1.2: If applicable, the geographic region and timeframe within which the study took place should be reported in the title or abstract.<br><br>RECORD 1.3: If linkage between databases was conducted for the study, this should be clearly stated in the title or abstract. | Title; Abstract (RECORD 1.1: CPRD Aurum named. RECORD 1.2: England, 2007-2024. RECORD 1.3: Linkage to HES and ONS stated in Abstract) |
| **Introduction** | | | | | |
| Background rationale | 2 | Explain the scientific background and rationale for the investigation being reported | Background | | Background |
| Objectives | 3 | State specific objectives, including any prespecified hypotheses | Background, final paragraph | | Background, final paragraph |
| **Methods** | | | | | |
| Study Design | 4 | Present key elements of study design early in the paper | Methods - Study design | | Methods - Study design |
| Setting | 5 | Describe the setting, locations, and relevant dates, including periods of recruitment, exposure, follow-up, and data collection | Methods - Study design | | Methods - Study design |
| Participants | 6 | *(a) Cohort study* - Give the eligibility criteria, and the sources and methods of selection of participants. Describe methods of follow-up<br>*Case-control study* - Give the eligibility criteria, and the sources and methods of case ascertainment and control selection. Give the rationale for the choice of cases | Methods - Participants | RECORD 6.1: The methods of study population selection (such as codes or algorithms used to identify subjects) should be listed in detail. If this is not possible, an explanation should be provided.<br><br>RECORD 6.2: Any validation studies of the codes or algorithms used to select the population should be referenced. If validation was conducted for this study and not | Methods - Participants; Procedures; Supplementary Figures 3a/3b (RECORD 6.1: infection SNOMED-CT codes; full lists at ref 28. RECORD 6.2: refs 25, 29, 30. RECORD 6.3: flow diagrams in Supplementary Figures 3a/3b) |

| | | | | | |
|---|---|---|---|---|---|
| | | and controls<br>*Cross-sectional study* - Give the eligibility criteria, and the sources and methods of selection of participants<br><br>*(b) Cohort study* - For matched studies, give matching criteria and number of exposed and unexposed<br>*Case-control study* - For matched studies, give matching criteria and the number of controls per case | | published elsewhere, detailed methods and results should be provided.<br><br>RECORD 6.3: If the study involved linkage of databases, consider use of a flow diagram or other graphical display to demonstrate the data linkage process, including the number of individuals with linked data at each stage. | |
| Variables | 7 | Clearly define all outcomes, exposures, predictors, potential confounders, and effect modifiers. Give diagnostic criteria, if applicable. | Methods - Procedures; Statistical analysis | RECORD 7.1: A complete list of codes and algorithms used to classify exposures, outcomes, confounders, and effect modifiers should be provided. If these cannot be reported, an explanation should be provided. | Methods - Procedures; Supplementary material (RECORD 7.1: outcome [self-harm SNOMED-CT; suicide ICD-10], exposure, and covariate codes fully published at ref 28) |
| Data sources/ measurement | 8 | For each variable of interest, give sources of data and details of methods of assessment (measurement).<br>Describe comparability of assessment methods if there is more than one group | Methods - Study design; Procedures; Supplementary Table 1 | | Methods - Study design; Procedures; Supplementary Table 1 |
| Bias | 9 | Describe any efforts to address potential sources of bias | Methods - Statistical analysis; Discussion - Limitations | | Methods - Statistical analysis; Discussion - Limitations |
| Study size | 10 | Explain how the study size was arrived at | Methods - Participants | | Methods - Participants |
| Quantitative variables | 11 | Explain how quantitative variables were handled in the analyses. If applicable, describe which groupings were chosen, and why | Methods - Procedures; Statistical analysis | | Methods - Procedures; Statistical analysis |
| Statistical methods | 12 | (a) Describe all statistical methods, including those used to control for confounding<br>(b) Describe any methods used to examine subgroups and interactions<br>(c) Explain how missing data were addressed<br>(d) *Cohort study* - If applicable, | Methods - Statistical analysis | | Methods - Statistical analysis |

| | | | | | |
|---|---|---|---|---|---|
| | | explain how loss to follow-up was addressed<br>*Case-control study* - If applicable, explain how matching of cases and controls was addressed<br>*Cross-sectional study* - If applicable, describe analytical methods taking account of sampling strategy<br>(e) Describe any sensitivity analyses | | | |
| Data access and cleaning methods | | .. | N/A (RECORD-only item) | RECORD 12.1: Authors should describe the extent to which the investigators had access to the database population used to create the study population.<br><br>RECORD 12.2: Authors should provide information on the data cleaning methods used in the study. | Methods - Study design; Data sharing statement (RECORD 12.1: patient-level access under CPRD licence ref 24_003746. RECORD 12.2: research-standard practices required; individuals with 1 day of follow-up excluded) |
| Linkage | | .. | N/A (RECORD-only item) | RECORD 12.3: State whether the study included person-level, institutional-level, or other data linkage across two or more databases. The methods of linkage and methods of linkage quality evaluation should be provided. | Methods - Study design (RECORD 12.3: person-level linkage of CPRD Aurum to HES APC and ONS mortality via NHS number, conducted by CPRD) |
| **Results** | | | | | |
| Participants | 13 | (a) Report the numbers of individuals at each stage of the study (*e.g.*, numbers potentially eligible, examined for eligibility, confirmed eligible, included in the study, completing follow-up, and analysed)<br>(b) Give reasons for non-participation at each stage.<br>(c) Consider use of a flow diagram | Results, paragraph 1; Supplementary Figures 3a/3b | RECORD 13.1: Describe in detail the selection of the persons included in the study (*i.e.,* study population selection) including filtering based on data quality, data availability and linkage. The selection of included persons can be described in the text and/or by means of the study flow diagram. | Results, paragraph 1; Supplementary Figures 3a/3b (RECORD 13.1: selection by ≥12 months registration, research-standard practice, and exclusion of 1-day follow-up; described in Methods and flow diagrams) |
| Descriptive data | 14 | (a) Give characteristics of study participants (*e.g.*, demographic, clinical, social) and information on exposures and potential confounders<br>(b) Indicate the number of participants with missing data for | Results; Table 1; Supplementary Table 2 | | Results; Table 1; Supplementary Table 2 |

| | | | | | |
|---|---|---|---|---|---|
| | | each variable of interest<br>(c) *Cohort study* - summarise follow-up time (*e.g.*, average and total amount) | | | |
| Outcome data | 15 | *Cohort study* - Report numbers of outcome events or summary measures over time<br>*Case-control study* - Report numbers in each exposure category, or summary measures of exposure<br>*Cross-sectional study* - Report numbers of outcome events or summary measures | Results; Supplementary Tables 3, 5 | | Results; Supplementary Tables 3, 5 |
| Main results | 16 | (a) Give unadjusted estimates and, if applicable, confounder-adjusted estimates and their precision (e.g., 95% confidence interval). Make clear which confounders were adjusted for and why they were included<br>(b) Report category boundaries when continuous variables were categorized<br>(c) If relevant, consider translating estimates of relative risk into absolute risk for a meaningful time period | Results; Figure 1; | | Results; Figure 1 |
| Other analyses | 17 | Report other analyses done-e.g., analyses of subgroups and interactions, and sensitivity analyses | Results; Tables 1-2; Supplementary Tables 3-6 | | Results; Tables 1-2; Supplementary Tables 3-6 |
| **Discussion** | | | | | |
| Key results | 18 | Summarise key results with reference to study objectives | Discussion, paragraph 1 | | Discussion, paragraph 1 |
| Limitations | 19 | Discuss limitations of the study, taking into account sources of potential bias or imprecision. Discuss both direction and magnitude of any potential bias | Discussion - Limitations | RECORD 19.1: Discuss the implications of using data that were not created or collected to answer the specific research question(s). Include discussion of misclassification bias, unmeasured confounding, missing data, and changing eligibility over time, as they pertain to the study being reported. | Discussion - Limitations (RECORD 19.1: misclassification of infection exposure [primary care = symptomatic cases], differential self-harm under-ascertainment, unmeasured confounding, missing covariate data, and changing eligibility via sequential matching all discussed) |
| Interpretation | 20 | Give a cautious overall interpretation of results considering | Discussion, final paragraph | | Discussion, final paragraph |

| | | | | | |
|---|---|---|---|---|---|
| | | objectives, limitations, multiplicity of analyses, results from similar studies, and other relevant evidence | | | |
| Generalisability | 21 | Discuss the generalisability (external validity) of the study results | Discussion - Limitations | | Discussion - Limitations |
| **Other Information** | | | | | |
| Funding | 22 | Give the source of funding and the role of the funders for the present study and, if applicable, for the original study on which the present article is based | Declaration of interests; Methods - Role of funding source | | |
| Accessibility of protocol, raw data, and programming code | | .. | | RECORD 22.1: Authors should provide information on how to access any supplemental information such as the study protocol, raw data, or programming code. | Data sharing statement; ref 28 at datacompass.lshtm.ac.uk (code lists). CPRD data via www.cprd.com application. Statistical analysis code not explicitly referenced. |

**Reference: Benchimol EI, Smeeth L, Guttmann A, Harron K, Moher D, Petersen I, Sørensen HT, von Elm E, Langan SM, the RECORD Working Committee. The REporting of studies Conducted using Observational Routinely-collected health Data (RECORD) Statement. PLoS Medicine 2015; in press.*

**Checklist is protected under Creative Commons Attribution (CC BY) license.*

#### Supplementary Table 1. Variable definitions

| Variable[1] | Category | Definition |
|---|---|---|
| **Exposures** | | |
| Gastroenteritis | Exposure | Primary care morbidity code (SNOMED-CT) for gastroenteritis, or symptom or pathogen code, recorded during the study period. |
| Lower respiratory tract infection (LRTI) | Exposure | Primary care morbidity code (SNOMED-CT) for LRTI recorded during the study period. |
| Skin/soft-tissue infection (SSTI) | Exposure | Primary care morbidity code (SNOMED-CT) for SSTI recorded during the study period. |
| Urinary tract infection (UTI) | Exposure | Primary care morbidity code (SNOMED-CT) for UTI recorded during the study period. |
| Sepsis | Exposure | Primary care morbidity code (SNOMED-CT) for sepsis recorded during the study period. |
| Meningitis/encephalitis | Exposure (positive control) | Primary care morbidity code (SNOMED-CT) for meningitis or encephalitis recorded during the study period. |
| **Outcomes** | | |
| Suicidal outcomes (composite) | Primary outcome | Earliest of: a first primary care diagnostic code for self-harm since index date (any act of self-poisoning or self-injury irrespective of intent) or a suicide death identified from Office for National Statistics (ONS) mortality records (ICD-10 coded). |
| Self-harm | Secondary outcome | First primary care diagnostic code for self-harm since index date, defined as any act of self-poisoning or self-injury irrespective of intent. |
| Suicide death | Secondary outcome | Suicide death identified from ONS mortality records using ICD-10 codes. |
| **Covariates** | | |
| Deprivation (IMD) | Covariate | Quintiles of individual-level Index of Multiple Deprivation (IMD), supplemented where missing with practice-level IMD data, and treated as a continuous ordinal score. |
| Harmful alcohol use | Covariate | Defined based on primary care morbidity codes suggesting harmful alcohol use (including alcohol dependency codes and codes related to physical/psychological harm from alcohol use) or a prescription for drugs used to maintain abstinence (acamprosate, disulfiram, or nalmefene). Individuals defined as harmful alcohol users if a relevant morbidity code or prescription was recorded on or before the index date. |
| Smoking status | Covariate | Defined pragmatically from primary care records using the status recorded closest to the index date, based on an algorithm where records identified within -1 year to +1 month of the index date were regarded as the best, +1 month to +1 year as second best, the nearest before -1 year as third best, and the nearest after +1 year as the worst. |

| Variable[1] | Category | Definition |
|---|---|---|
| Obesity | Covariate | Defined as BMI ≥30 kg/m², based on primary care records. BMI was calculated using height and weight measures recorded closest to the index date (prioritising the period from 1 year before to 1 month after the index date; if unavailable, the nearest value in the year following the index date was used; if still unavailable, the nearest value in the year before the index date was used; if still unavailable, the nearest weight recorded beyond one year after the index date was used). Where a calculated BMI was unavailable, directly recorded BMI values were used. Treated as missing if unrecorded.(1) |
| Charlson Comorbidity Index (CCI) | Covariate | Calculated from primary care morbidity coding. Comorbidities and weights: myocardial infarction, congestive heart failure, peripheral vascular disease, cerebrovascular disease, dementia, lung disease, rheumatoid diseases, peptic ulcer, mild liver disease, diabetes without complications (weight 1); diabetes with complications, hemiplegia, chronic kidney disease, cancer including lymphoma (weight 2); moderate/severe liver disease (weight 3); metastatic cancer and HIV (weight 6). Categorised as low (0), moderate (1-2), or severe (≥3).(2) |
| Common mental disorders (CMD) | Covariate | Pre-index primary care morbidity code for depression or anxiety disorder. |
| Severe mental illness (SMI) | Covariate | Pre-index primary care morbidity code for bipolar disorder, schizophrenia, or other psychosis. |
| Prior self-harm | Covariate | Any primary care record of self-harm prior to the index date, based on morbidity coding. |
| Recreational injecting drug use | Covariate (SSTI cohort only) | Primary care morbidity code for recreational injecting drug use recorded on or before the index date. |
| Ethnicity | Covariate | Identified from primary care records using a previously validated algorithm.(3) |
| Healthcare utilisation | Covariate | Number of primary care consultations in the 12 months prior to the index date. |
| Infection severity | Effect modifier / secondary exposure | Three-level exposure among participants with HES linkage: no infection; non-severe infection (CPRD primary care record only); severe infection (CPRD record plus HES record of hospital admission for the same infection or for sepsis within 28 days either side of the CPRD record). Assessed in gastroenteritis, LRTI, SSTI, and UTI cohorts only. |
| **Effect modifiers/secondary exposures** | | |
| Antimicrobial prescription | Effect modifier / secondary exposure | Three-level exposure: unexposed; untreated infection (no antimicrobial prescription at index date); treated infection (antimicrobial prescription recorded within 7 days of index date). Drug classes included: penicillins, fluoroquinolones, macrolides, and cephalosporins, plus drugs listed in the NICE BNF treatment summary for UTIs. Assessed in gastroenteritis, LRTI, SSTI, and UTI cohorts only. |
| Frailty | Effect modifier | Assessed using the electronic Frailty Index (eFI) Version 1, which uses primary care morbidity coding across 36 deficits to categorise individuals as fit, mildly frail, moderately frail, or severely frail. Analysis restricted to participants aged ≥65 years, the age group in which the eFI has been validated. |

| Variable[1] | Category | Definition |
|---|---|---|
| Prior mental health conditions | Effect modifier | Composite binary variable: any pre-index record of CMD (depression, anxiety), SMI (bipolar disorder, schizophrenia, other psychosis), or self-harm. |
| Age | Effect modifier | Categorised as 18-29, 30-39, 40-49, 50-64, and ≥65 years based on age at cohort entry. |
| Sex | Effect modifier | Binary variable (female/male) based on CPRD general practice registration records. |

*CMD: common mental disorders; CCI: Charlson Comorbidity Index; CPRD: Clinical Practice Research Datalink; eFI: electronic Frailty Index; HES: Hospital Episode Statistics; IMD: Index of Multiple Deprivation; LRTI: lower respiratory tract infection; ONS: Office for National Statistics; SMI: severe mental illness; SSTI: skin/soft-tissue infection; UTI: urinary tract infection.*

**Supplementary Table 2. Missing data summary and complete case comparison (figures are n [%] unless otherwise specified)**

| Variable | | Included in CC analysis | Excluded (missing covariates) |
|---|---|---|---|
| **Meningitis/Encephalitis** | | | |
| **Total** | | 96,585 | 12,522 |
| **Age, median [IQR]** | | 45.00 [32.00, 62.00] | 29.00 [22.00, 41.00] |
| **Age category (%)** | 18-29 | 19,519 (20.21) | 6,347 (50.69) |
| | 30-39 | 20,403 (21.12) | 2,865 (22.88) |
| | 40-49 | 15,654 (16.21) | 1,281 (10.23) |
| | 50-64 | 20,495 (21.22) | 1,211 (9.67) |
| | 65+ | 20,514 (21.24) | 818 (6.53) |
| **Sex (%)** | Female | 56,765 (58.77) | 4,288 (34.24) |
| | Male | 39,820 (41.23) | 8,234 (65.76) |
| **Ethnicity (%)** | White | 62,246 (73.66) | 5,249 (75.80) |
| | South Asian | 4,162 (4.92) | 446 (6.44) |
| | Black | 2,012 (2.38) | 222 (3.21) |
| | Mixed | 15,531 (18.38) | 902 (13.03) |
| | Other | 558 (0.66) | 106 (1.53) |
| | Missing | 12,076 (12.50) | 5,597 (44.70) |
| **IMD (%)** | 1, least deprived | 19,754 (20.45) | 2,434 (20.03) |
| | 2 | 18,418 (19.07) | 2,259 (18.59) |
| | 3 | 17,905 (18.54) | 2,256 (18.57) |
| | 4 | 20,526 (21.25) | 2,639 (21.72) |
| | 5, most deprived | 19,982 (20.69) | 2,562 (21.09) |
| | Missing | 0 (0.00) | 372 (2.97) |
| **Harmful alcohol use (%)** | Yes | 1,998 (2.07) | 150 (1.20) |
| **Smoking status (%)** | Non-smoker | 52,553 (54.41) | 5,334 (56.82) |
| | Current- or ex-smoker | 44,032 (45.59) | 4,054 (43.18) |
| | NA | 0 (0.00) | 3,134 (25.03) |
| **Obesity (%)** | Not obese | 73,316 (75.91) | 648 (80.00) |

| Variable | | Included in CC analysis | Excluded (missing covariates) |
|---|---|---|---|
| | Obese | 23,269 (24.09) | 162 (20.00) |
| | NA | 0 (0.00) | 11,712 (93.53) |
| **Number of consultations in year prior, median [IQR]** | | 10.00 [4.00, 20.00] | 3.00 [1.00, 8.00] |
| **CCI category (%)** | Low (0) | 59,939 (62.06) | 10,573 (84.44) |
| | Moderate (1-2) | 26,451 (27.39) | 1,717 (13.71) |
| | Severe (3 or more) | 10,195 (10.56) | 232 (1.85) |
| **Gastroenteritis** | | | |
| **Total** | | 1,537,301 | 180,487 |
| **Age, median [IQR]** | | 52.00 [35.00, 70.00] | 32.00 [23.00, 52.00] |
| **Age category (%)** | 18-29 | 253,176 (16.47) | 80,099 (44.38) |
| | 30-39 | 232,371 (15.12) | 32,119 (17.80) |
| | 40-49 | 221,510 (14.41) | 19,297 (10.69) |
| | 50-64 | 336,876 (21.91) | 19,534 (10.82) |
| | 65+ | 493,368 (32.09) | 29,438 (16.31) |
| **Sex (%)** | Female | 913,065 (59.39) | 71,808 (39.79) |
| | Male | 624,236 (40.61) | 108,679 (60.21) |
| **Ethnicity (%)** | White | 977,243 (73.59) | 72,712 (74.55) |
| | South Asian | 65,255 (4.91) | 6,952 (7.13) |
| | Black | 30,806 (2.32) | 3,505 (3.59) |
| | Mixed | 246,623 (18.57) | 12,994 (13.32) |
| | Other | 8,019 (0.60) | 1,367 (1.40) |
| | NA | 209,355 (13.62) | 82,957 (45.96) |
| **IMD (%)** | 1, least deprived | 313,965 (20.42) | 34,327 (19.61) |
| | 2 | 292,936 (19.06) | 31,361 (17.91) |
| | 3 | 290,845 (18.92) | 33,268 (19.00) |
| | 4 | 326,440 (21.23) | 39,083 (22.32) |
| | 5, most deprived | 313,115 (20.37) | 37,041 (21.16) |
| | NA | 0 (0.00) | 5,407 (3.00) |

| Variable | | Included in CC analysis | Excluded (missing covariates) |
|---|---|---|---|
| **Harmful alcohol use (%)** | Yes | 29,295 (1.91) | 2,098 (1.16) |
| **Smoking status (%)** | Non-smoker | 834,298 (54.27) | 78,550 (57.57) |
| | Current- or ex-smoker | 703,003 (45.73) | 57,901 (42.43) |
| | NA | 0 (0.00) | 44,036 (24.40) |
| **Obesity (%)** | Not obese | 1,172,721 (76.28) | 8,980 (80.16) |
| | Obese | 364,580 (23.72) | 2,223 (19.84) |
| | NA | 0 (0.00) | 169,284 (93.79) |
| **Number of consultations in year prior, median [IQR]** | | 11.00 [5.00, 20.00] | 3.00 [1.00, 8.00] |
| **CCI category (%)** | Low (0) | 893,678 (58.13) | 146,076 (80.93) |
| | Moderate (1-2) | 435,913 (28.36) | 27,300 (15.13) |
| | Severe (3 or more) | 207,710 (13.51) | 7,111 (3.94) |
| **LRTI** | | | |
| **Total** | | 1,826,609 | 181,344 |
| **Age, median [IQR]** | | 57.00 [42.00, 71.00] | 42.00 [27.00, 65.00] |
| **Age category (%)** | 18-29 | 168,869 (9.24) | 52,883 (29.16) |
| | 30-39 | 232,701 (12.74) | 31,094 (17.15) |
| | 40-49 | 282,478 (15.46) | 24,378 (13.44) |
| | 50-64 | 472,087 (25.84) | 27,609 (15.22) |
| | 65+ | 670,474 (36.71) | 45,380 (25.02) |
| **Sex (%)** | Female | 1,051,307 (57.56) | 75,036 (41.38) |
| | Male | 775,302 (42.44) | 106,308 (58.62) |
| **Ethnicity (%)** | White | 1,187,090 (75.10) | 73,990 (77.92) |
| | South Asian | 68,725 (4.35) | 5,625 (5.92) |
| | Black | 30,825 (1.95) | 2,560 (2.70) |
| | Mixed | 286,581 (18.13) | 11,739 (12.36) |
| | Other | 7,361 (0.47) | 1,040 (1.10) |
| | NA | 246,027 (13.47) | 86,390 (47.64) |
| **IMD (%)** | 1, least deprived | 371,218 (20.32) | 34,039 (19.61) |

| Variable | | Included in CC analysis | Excluded (missing covariates) |
|---|---|---|---|
| | 2 | 354,087 (19.38) | 31,624 (18.22) |
| | 3 | 337,083 (18.45) | 32,531 (18.74) |
| | 4 | 380,783 (20.85) | 37,845 (21.80) |
| | 5, most deprived | 383,438 (20.99) | 37,563 (21.64) |
| | NA | 0 (0.00) | 7,742 (4.27) |
| **Harmful alcohol use (%)** | Yes | 37,435 (2.05) | 2,519 (1.39) |
| **Smoking status (%)** | Non-smoker | 968,830 (53.04) | 78,722 (56.68) |
| | Current- or ex-smoker | 857,779 (46.96) | 60,175 (43.32) |
| | NA | 0 (0.00) | 42,447 (23.41) |
| **Obesity (%)** | Not obese | 1,373,141 (75.17) | 9,481 (77.94) |
| | Obese | 453,468 (24.83) | 2,683 (22.06) |
| | NA | 0 (0.00) | 169,180 (93.29) |
| **Number of consultations in year prior, median [IQR]** | | 11.00 [5.00, 20.00] | 3.00 [1.00, 10.00] |
| **CCI category (%)** | Low (0) | 1,015,345 (55.59) | 138,999 (76.65) |
| | Moderate (1-2) | 543,358 (29.75) | 31,793 (17.53) |
| | Severe (3 or more) | 267,906 (14.67) | 10,552 (5.82) |
| **SSTI** | | | |
| **Total** | | 1,919,299 | 222,547 |
| **Age, median [IQR]** | | 52.00 [37.00, 68.00] | 33.00 [23.00, 52.00] |
| **Age category (%)** | 18-29 | 276,274 (14.39) | 93,784 (42.14) |
| | 30-39 | 291,044 (15.16) | 39,915 (17.94) |
| | 40-49 | 315,069 (16.42) | 27,794 (12.49) |
| | 50-64 | 457,068 (23.81) | 26,648 (11.97) |
| | 65+ | 579,844 (30.21) | 34,406 (15.46) |
| **Sex (%)** | Female | 1,108,169 (57.74) | 83,739 (37.63) |
| | Male | 811,130 (42.26) | 138,808 (62.37) |
| **Ethnicity (%)** | White | 1,237,279 (74.20) | 91,734 (75.56) |
| | South Asian | 79,297 (4.76) | 8,253 (6.80) |

| Variable | | Included in CC analysis | Excluded (missing covariates) |
|---|---|---|---|
| | Black | 35,862 (2.15) | 4,088 (3.37) |
| | Mixed | 305,578 (18.33) | 15,658 (12.90) |
| | Other | 9,460 (0.57) | 1,670 (1.38) |
| | NA | 251,823 (13.12) | 101,144 (45.45) |
| **IMD (%)** | 1, least deprived | 405,009 (21.10) | 43,567 (20.12) |
| | 2 | 375,986 (19.59) | 39,817 (18.39) |
| | 3 | 360,462 (18.78) | 40,954 (18.91) |
| | 4 | 399,628 (20.82) | 47,605 (21.98) |
| | 5, most deprived | 378,214 (19.71) | 44,598 (20.60) |
| | NA | 0 (0.00) | 6,006 (2.70) |
| **Harmful alcohol use (%)** | Yes | 37,157 (1.94) | 2,698 (1.21) |
| **Smoking status (%)** | Non-smoker | 1,037,302 (54.05) | 95,257 (57.24) |
| | Current- or ex-smoker | 881,997 (45.95) | 71,165 (42.76) |
| | NA | 0 (0.00) | 56,125 (25.22) |
| **Injectable drug use (%)** | Yes | 9,457 (0.49) | 1,011 (0.45) |
| **Obesity (%)** | Not obese | 1,456,593 (75.89) | 10,634 (79.73) |
| | Obese | 462,706 (24.11) | 2,703 (20.27) |
| | NA | 0 (0.00) | 209,210 (94.01) |
| **Number of consultations in year prior, median [IQR]** | | 10.00 [4.00, 20.00] | 3.00 [1.00, 8.00] |
| **CCI category (%)** | Low (0) | 1,138,393 (59.31) | 181,322 (81.48) |
| | Moderate (1-2) | 541,105 (28.19) | 33,073 (14.86) |
| | Severe (3 or more) | 239,801 (12.49) | 8,152 (3.66) |
| **UTI** | | | |
| **Total** | | 986,009 | 81,214 |
| **Age, median [IQR]** | | 56.00 [36.00, 74.00] | 41.00 [24.00, 73.00] |
| **Age category (%)** | 18-29 | 158,281 (16.05) | 28,462 (35.05) |
| | 30-39 | 129,563 (13.14) | 10,998 (13.54) |
| | 40-49 | 122,054 (12.38) | 7,603 (9.36) |

| Variable | | Included in CC analysis | Excluded (missing covariates) |
|---|---|---|---|
| | 50-64 | 196,090 (19.89) | 9,369 (11.54) |
| | 65+ | 380,021 (38.54) | 24,782 (30.51) |
| **Sex (%)** | Female | 783,495 (79.46) | 63,387 (78.05) |
| | Male | 202,514 (20.54) | 17,827 (21.95) |
| **Ethnicity (%)** | White | 635,546 (74.89) | 33,056 (75.21) |
| | South Asian | 36,309 (4.28) | 2,766 (6.29) |
| | Black | 17,132 (2.02) | 1,520 (3.46) |
| | Mixed | 155,017 (18.27) | 6,034 (13.73) |
| | Other | 4,591 (0.54) | 575 (1.31) |
| | NA | 137,414 (13.94) | 37,263 (45.88) |
| **IMD (%)** | 1, least deprived | 209,329 (21.23) | 16,259 (20.94) |
| | 2 | 192,938 (19.57) | 14,439 (18.59) |
| | 3 | 184,640 (18.73) | 14,693 (18.92) |
| | 4 | 205,930 (20.89) | 17,114 (22.04) |
| | 5, most deprived | 193,172 (19.59) | 15,157 (19.52) |
| | NA | 0 (0.00) | 3,552 (4.37) |
| **Alcohol abuse (%)** | Yes | 14,577 (1.48) | 829 (1.02) |
| **Smoking status (%)** | Non-smoker | 558,960 (56.69) | 40,038 (62.60) |
| | Current- or ex-smoker | 427,049 (43.31) | 23,924 (37.40) |
| | NA | 0 (0.00) | 17,252 (21.24) |
| **Obesity (%)** | Not obese | 752,503 (76.32) | 4,590 (77.81) |
| | Obese | 233,506 (23.68) | 1,309 (22.19) |
| | NA | 0 (0.00) | 75,315 (92.74) |
| **Number of consultations in year prior, median [IQR]** | | 12.00 [6.00, 22.00] | 5.00 [1.00, 12.00] |
| **CCI category (%)** | Low (0) | 544,546 (55.23) | 60,081 (73.98) |
| | Moderate (1-2) | 286,512 (29.06) | 15,154 (18.66) |
| | Severe (3 or more) | 154,951 (15.71) | 5,979 (7.36) |
| **Sepsis** | | | |

| Variable | | Included in CC analysis | Excluded (missing covariates) |
|---|---|---|---|
| **Total** | | 1,074,978 | 63,886 |
| **Age, median [IQR]** | | 73.00 [60.00, 82.00] | 66.00 [41.00, 83.00] |
| **Age category (%)** | 18-29 | 32,014 (2.98) | 9,091 (14.23) |
| | 30-39 | 47,172 (4.39) | 5,952 (9.32) |
| | 40-49 | 67,391 (6.27) | 5,349 (8.37) |
| | 50-64 | 199,470 (18.56) | 10,495 (16.43) |
| | 65+ | 728,931 (67.81) | 32,999 (51.65) |
| **Sex (%)** | Female | 530,348 (49.34) | 28,359 (44.39) |
| | Male | 544,630 (50.66) | 35,527 (55.61) |
| **Ethnicity (%)** | White | 730,651 (75.95) | 28,794 (80.02) |
| | South Asian | 32,686 (3.40) | 1,742 (4.84) |
| | Black | 15,592 (1.62) | 891 (2.48) |
| | Mixed | 180,130 (18.72) | 4,250 (11.81) |
| | Other | 2,971 (0.31) | 307 (0.85) |
| | NA | 112,948 (10.51) | 27,902 (43.67) |
| **IMD (%)** | 1, least deprived | 215,984 (20.09) | 11,775 (19.49) |
| | 2 | 210,297 (19.56) | 11,003 (18.21) |
| | 3 | 200,947 (18.69) | 11,659 (19.30) |
| | 4 | 226,396 (21.06) | 13,333 (22.07) |
| | 5, most deprived | 221,354 (20.59) | 12,646 (20.93) |
| | NA | 0 (0.00) | 3,470 (5.43) |
| **Alcohol abuse (%)** | Yes | 26,685 (2.48) | 1,142 (1.79) |
| **Smoking status (%)** | Non-smoker | 545,684 (50.76) | 29,042 (56.74) |
| | Current- or ex-smoker | 529,294 (49.24) | 22,141 (43.26) |
| | NA | 0 (0.00) | 12,703 (19.88) |
| **Obesity (%)** | Not obese | 794,724 (73.93) | 3,757 (75.65) |
| | Obese | 280,254 (26.07) | 1,209 (24.35) |
| | NA | 0 (0.00) | 58,920 (92.23) |

| Variable | | Included in CC analysis | Excluded (missing covariates) |
|---|---|---|---|
| **Number of consultations in year prior, median [IQR]** | | 18.00 [9.00, 31.00] | 7.00 [2.00, 19.00] |
| **CCI category (%)** | Low (0) | 368,478 (34.28) | 38,914 (60.91) |
| | Moderate (1-2) | 352,938 (32.83) | 15,519 (24.29) |
| | Severe (3 or more) | 353,562 (32.89) | 9,453 (14.80) |

*Complete-case analysis restricted to participants with complete data on all Model 3 covariates (IMD, alcohol use, smoking status, obesity, CCI score, prior CMD, prior SMI, prior self-harm; injectable drug use additionally required for SSTI cohort). Excluded: participants in the post-exclusion matched cohort who were not retained after complete-case restriction due to missing data in ≥ 1 covariate. N (%) shown; percentages calculated among non-missing values unless stated. IMD: Index of Multiple Deprivation; CCI: Charlson Comorbidity Index; CMD: common mental disorders; SMI: severe mental illness.*

**Supplementary Table 3. Model-derived rate differences (per 1,000 person-years) between infected and uninfected participants (full follow-up period)**

| Infection | Events (infected) | Person-years (infected) | Observed rate (per 1,000 PY) | Model 3 HR | Counterfactual rate (per 1,000 PY) | RD (95% CI) (per 1,000 PY) |
|---|---|---|---|---|---|---|
| Gastroenteritis | 5,127 | 1,939,099.0 | 2.64 | 1.620 | 1.63 | 1.01 (0.93 to 1.09) |
| LRTI | 5,033 | 2,491,828.0 | 2.02 | 1.369 | 1.48 | 0.54 (0.47 to 0.62) |
| Meningitis/Encephalitis | 351 | 103,385.0 | 3.40 | 1.560 | 2.18 | 1.22 (0.84 to 1.60) |
| Sepsis | 1,901 | 583,900.8 | 3.26 | 1.786 | 1.82 | 1.43 (1.28 to 1.59) |
| SSTI | 5,403 | 2,584,921.8 | 2.09 | 1.368 | 1.53 | 0.56 (0.49 to 0.63) |
| UTI | 2,641 | 1,208,245.4 | 2.19 | 1.411 | 1.55 | 0.64 (0.54 to 0.73) |

*PY: person-years. HR: hazard ratio from Model 3 (fully adjusted Cox model), fitted on the full follow-up period. Observed rate: events / person-years x 1,000 in the infected group of the complete-case analytic sample. Counterfactual rate: observed infected rate x (1 / HR), representing the expected rate had the infected group been uninfected. RD: rate difference (observed minus counterfactual), per 1,000 person-years. 95% CI derived via delta method, propagating Poisson uncertainty in the observed rate and log-normal uncertainty in the HR.*

#### Supplementary Table 4. Sensitivity analyses results

| | Analysis | Description/justification | Meningitis/Encephalitis | Gastroenteritis | LRTI | SSTI | UTI | Sepsis |
|---|---|---|---|---|---|---|---|---|
| **Main** | **Main analysis (fully adjusted)** | **Included for comparison.** | **1.56 (1.32-1.84)** | **1.62 (1.55-1.70)** | **1.37 (1.31-1.44)** | **1.37 (1.31-1.43)** | **1.41 (1.33-1.50)** | **1.79 (1.65-1.93)** |
| 1 | Pre-COVID (end 1 March 2020) | Repeat analysis ending 1 March 2020 to assess pandemic impact. | 1.73 (1.42-2.11) | 1.61 (1.52-1.70) | 1.34 (1.27-1.42) | 1.32 (1.25-1.39) | 1.48 (1.38-1.59) | 1.68 (1.52-1.87) |
| 2 | 1-year lagged start | Follow-up starts 1 year post-infection; CMD events in year 1 excluded. | 1.39 (1.15-1.69) | 1.54 (1.46-1.63) | 1.37 (1.30-1.44) | 1.34 (1.27-1.41) | 1.34 (1.25-1.43) | 1.66 (1.51-1.83) |
| 3 | Active patients only (>=1 GP consultation prior year) | Excludes practice non-attenders. | 1.53 (1.29-1.81) | 1.58 (1.51-1.66) | 1.33 (1.27-1.40) | 1.33 (1.27-1.39) | 1.38 (1.30-1.47) | 1.75 (1.62-1.89) |
| 4 | Missing ethnicity as 'Unknown' category | Investigates effect of missing ethnicity data. | 1.56 (1.32-1.84) | 1.63 (1.55-1.70) | 1.37 (1.31-1.44) | 1.37 (1.31-1.43) | 1.41 (1.33-1.50) | 1.78 (1.65-1.93) |
| 5 | Exclude self-harm in prior 2 years | Most recent self-harm within 2 years of index date excluded. | 1.58 (1.33-1.87) | 1.64 (1.56-1.72) | 1.38 (1.31-1.45) | 1.37 (1.30-1.43) | 1.40 (1.31-1.49) | 1.78 (1.64-1.92) |
| 6 | Suicide/self-harm (individual outcome) | Primary analysis repeated for suicide/self-harm as outcome. | 3.13 (0.92-10.71) | 1.24 (0.94-1.63) | 1.06 (0.82-1.38) | 1.07 (0.83-1.39) | 1.83 (1.22-2.74) | 1.84 (1.20-2.80) |
| 7 | Any self-harm (individual outcome) | Primary analysis repeated for any self-harm as outcome. | 1.53 (1.29-1.81) | 1.63 (1.56-1.71) | 1.38 (1.32-1.45) | 1.38 (1.32-1.44) | 1.40 (1.32-1.49) | 1.79 (1.66-1.94) |

*HR: hazard ratio; CI: confidence interval. N: total participants (exposed); events: outcome events. All models fully adjusted (Model 3). GE: gastroenteritis; LRTI: lower respiratory tract infection; SSTI: skin and soft tissue infection; UTI: urinary tract infection.*

**Supplementary Table 5. Association between infection and suicide/self-harm across cumulative follow-up windows**

| Dataset | Follow-up band (years) | Events | HR (95% CI)[1] | P value[2] |
|---|---|---|---|---|
| Meningitis/Encephalitis | 0-1 | 276 | 2.20 (1.55-3.12) | <0.0001 |
| | 0-2 | 468 | 1.96 (1.52-2.55) | |
| | 0-3 | 588 | 1.77 (1.41-2.23) | |
| | 0-4 | 711 | 1.62 (1.32-2.00) | |
| | 0-5 | 802 | 1.62 (1.33-1.97) | |
| Gastroenteritis | 0-1 | 3,641 | 1.89 (1.72-2.08) | |
| | 0-2 | 5,849 | 1.81 (1.68-1.95) | |
| | 0-3 | 7,455 | 1.75 (1.64-1.87) | |
| | 0-4 | 8,812 | 1.71 (1.61-1.82) | |
| | 0-5 | 9,860 | 1.67 (1.58-1.76) | |
| LRTI | 0-1 | 3,521 | 1.34 (1.21-1.47) | |
| | 0-2 | 5,821 | 1.42 (1.32-1.53) | |
| | 0-3 | 7,500 | 1.40 (1.31-1.50) | |
| | 0-4 | 8,912 | 1.40 (1.31-1.48) | |
| | 0-5 | 9,979 | 1.39 (1.32-1.47) | |
| SSTI | 0-1 | 4,218 | 1.46 (1.33-1.59) | |
| | 0-2 | 6,830 | 1.41 (1.32-1.51) | |
| | 0-3 | 8,810 | 1.38 (1.30-1.46) | |
| | 0-4 | 10,435 | 1.38 (1.31-1.46) | |
| | 0-5 | 11,623 | 1.39 (1.32-1.47) | |
| UTI | 0-1 | 2,183 | 1.66 (1.47-1.88) | |
| | 0-2 | 3,552 | 1.49 (1.36-1.64) | |
| | 0-3 | 4,564 | 1.43 (1.31-1.56) | |
| | 0-4 | 5,404 | 1.44 (1.33-1.56) | |
| | 0-5 | 6,121 | 1.43 (1.33-1.54) | |
| Sepsis | 0-1 | 1,845 | 1.98 (1.72-2.27) | |
| | 0-2 | 2,898 | 1.91 (1.71-2.14) | |
| | 0-3 | 3,772 | 1.83 (1.66-2.02) | |

| Dataset | Follow-up band (years) | Events | HR (95% CI)[1] | P value[2] |
|---|---|---|---|---|
| | 0-4 | 4,440 | 1.79 (1.63-1.96) | |
| | 0-5 | 4,932 | 1.78 (1.63-1.94) | |

*HR: hazard ratio; CI: confidence interval. LRTI: lower respiratory tract infections; SSTI: skin and soft tissue infections; UTI: urinary tract infections*

*[1]Fully adjusted for deprivation, harmful alcohol use, smoking status, obesity, Charlson Comorbidity Index score, prior common mental disorders, severe mental illness, and pre-index self-harm*

*[2]From a likelihood ratio test comparing the model with the main exposure effect against one including a time-interaction term*

**Supplementary Table 6. Model-derived rate differences (per 1,000 person-years) between infected and uninfected participants (first-year post-infection)**

| Infection | Events (infected) | Person-years (infected) | Observed rate (per 1,000 PY) | Model 3 HR | Counterfactual rate (per 1,000 PY) | RD (95% CI) (per 1,000 PY) |
|---|---|---|---|---|---|---|
| Gastroenteritis | 1,426 | 272,774.8 | 5.23 | 1.888 | 2.77 | 2.46 (2.17 to 2.75) |
| LRTI | 1,211 | 336,822.6 | 3.60 | 1.362 | 2.64 | 0.95 (0.69 to 1.22) |
| Meningitis/Encephalitis | 108 | 15,575.2 | 6.93 | 2.196 | 3.16 | 3.78 (2.47 to 5.09) |
| Sepsis | 722 | 140,689.6 | 5.13 | 2.066 | 2.48 | 2.65 (2.26 to 3.04) |
| SSTI | 1,366 | 354,540.8 | 3.85 | 1.457 | 2.65 | 1.21 (0.96 to 1.45) |
| UTI | 710 | 170,285.1 | 4.17 | 1.669 | 2.50 | 1.67 (1.34 to 2.00) |

*PY = person-years. HR = hazard ratio from Model 3 (fully adjusted Cox model), fitted on the 0–1 year post-infection. Observed rate = events / person-years x 1,000 in the infected group of the complete-case analytic sample. Counterfactual rate = observed infected rate x (1 / HR), representing the expected rate had the infected group been uninfected. RD = rate difference (observed minus counterfactual), per 1,000 person-years. 95% CI derived via delta method, propagating Poisson uncertainty in the observed rate and log-normal uncertainty in the HR.*

#### Supplementary Table 7. Additional variable information

| | | Meningitis/ Encephalitis | | Gastroenteritis | | LRTI | | SSTI | | UTI | | Sepsis | |
|---|---|---|---|---|---|---|---|---|---|---|---|---|---|
| **Variable[1]** | **Level** | **Matched comparators** | **With infection** | **Matched comparators** | **With infection** | **Matched comparators** | **With infection** | **Variable[1]** | **Level** | **Matched comparators** | **With infection** | **Matched comparators** | **With infection** |
| **Total** | | 90,915 | 18,192 | 1,405,871 | 311,917 | 1,627,978 | 379,975 | **Total** | | 90,915 | 18,192 | 1,405,871 | 311,917 |
| **Injectable drug use (%)** | Yes | - | - | - | - | - | - | 7,344 (0.42) | 3,124 (0.78) | - | - | - | - |
| **Frailty category (%)** | Fit (≤0.12) | 76,462 (84.10) | 13,599 (74.75) | 1,111,496 (79.06) | 208,357 (66.80) | 1,254,253 (77.04) | 238,070 (62.65) | 1,403,484 (80.49) | 278,944 (70.07) | 644,321 (73.66) | 116,151 (60.33) | 487,499 (51.43) | 55,979 (29.32) |
| | Mild frailty (0.13-0.24) | 10,145 (11.16) | 3,028 (16.64) | 198,181 (14.10) | 63,093 (20.23) | 256,311 (15.74) | 88,376 (23.26) | 233,233 (13.38) | 76,276 (19.16) | 147,837 (16.90) | 43,750 (22.72) | 250,814 (26.46) | 55,179 (28.90) |
| | Moderate frailty (0.25-0.36) | 3,155 (3.47) | 1,140 (6.27) | 71,692 (5.10) | 28,019 (8.98) | 88,804 (5.45) | 37,763 (9.94) | 80,132 (4.60) | 30,477 (7.66) | 60,255 (6.89) | 21,625 (11.23) | 138,304 (14.59) | 44,801 (23.47) |
| | Severe frailty (>0.37) | 1,153 (1.27) | 425 (2.34) | 24,502 (1.74) | 12,448 (3.99) | 28,610 (1.76) | 15,766 (4.15) | 26,898 (1.54) | 12,402 (3.12) | 22,290 (2.55) | 10,994 (5.71) | 71,334 (7.53) | 34,954 (18.31) |
| **Antimicrobial prescription on index date (%)** | Yes | 0 (0.00) | 234 (1.29) | 0 (0.00) | 16,372 (5.25) | 0 (0.00) | 267,351 (70.36) | 0 (0.00) | 238,671 (59.95) | 0 (0.00) | 130,268 (67.66) | 0 (0.00) | 5,867 (3.07) |
| **Severe infection (%)** | Yes | — | 7,113 (39.10) | — | 4,899 (1.57) | — | 21,910 (5.77) | — | 11,381 (2.86) | — | 11,512 (5.98) | — | 60,605 (31.74) |

| | | | | | | | | | | | | |
|---|---|---|---|---|---|---|---|---|---|---|---|---|
| **Time to event (events only), median[3] [IQR]** | 3.09 [1.12, 5.90] | 1.98 [0.73, 4.69] | 2.75 [1.01, 5.76] | 2.46 [0.82, 5.55] | 2.74 [1.00, 5.75] | 2.79 [1.05, 5.99] | 2.58 [0.93, 5.49] | 2.65 [0.97, 5.77] | 2.79 [1.06, 5.86] | 2.72 [0.91, 5.72] | 2.50 [0.94, 4.78] | 1.67 [0.54, 3.62] |

#### Supplementary Figure 1. Study design diagram

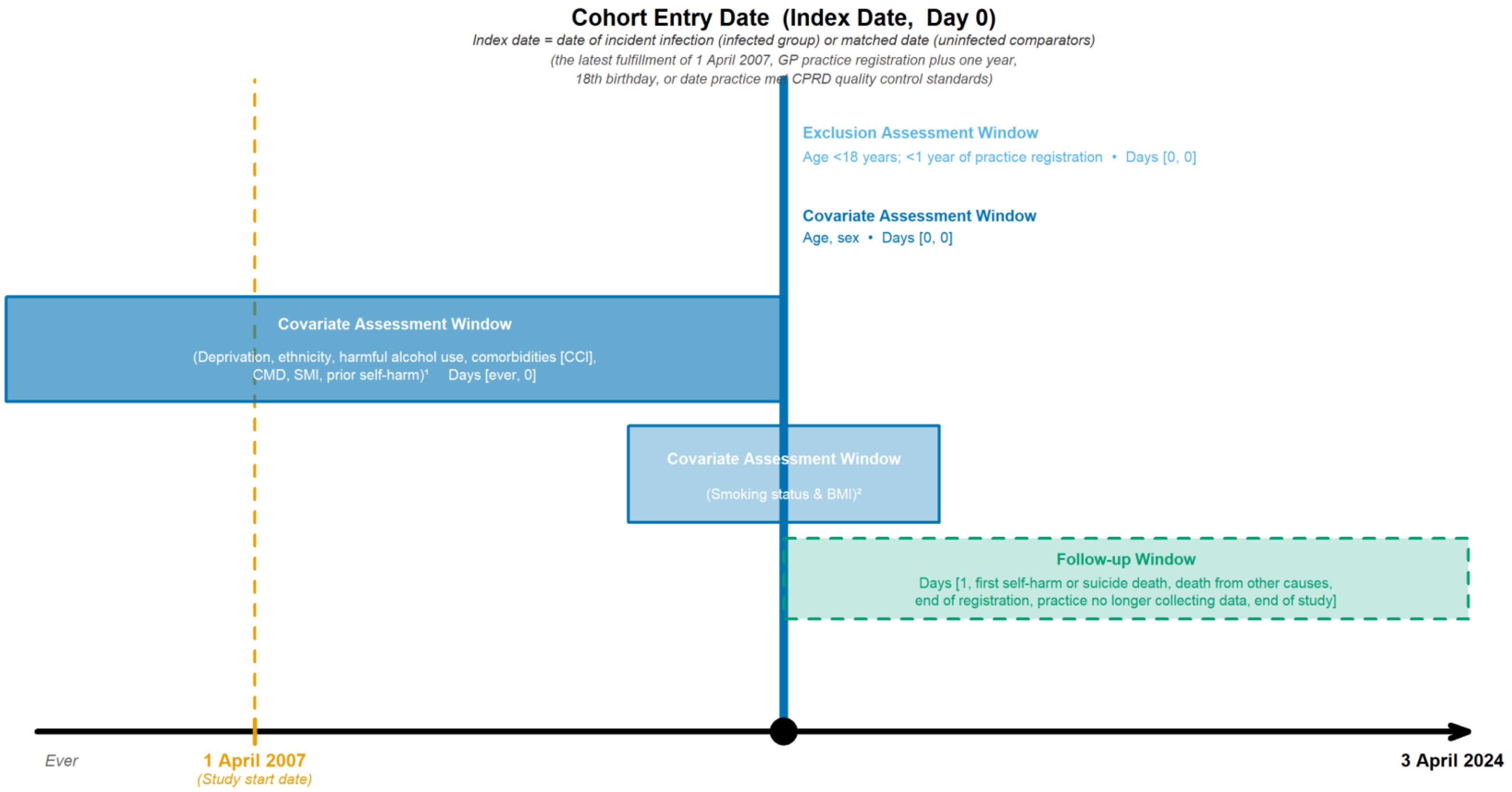


*[1] Injecting drug use assessed as covariate in skin & soft tissue infection (SSTI) cohort only.*

*[2] Smoking status and BMI identified using an algorithm where records within -1 year to +1 month of the index date are regarded as best, +1 month to +1 year from index date as second best, the nearest before -1 year as third best, and the nearest after +1 year as worst.*

*CCI = Charlson Comorbidity Index. CMD = common mental disorder. SMI = severe mental illness. IDU = injecting drug use.*

**Supplementary Figure 2. Directed acyclic graph illustrating hypothesized confounding relationships between infection and suicide or self-harm**

**Supplementary Figure 1. Directed acyclic graph of hypothesised confounding relationships**

Arrows represent hypothesised causal paths; confounders shown in colour

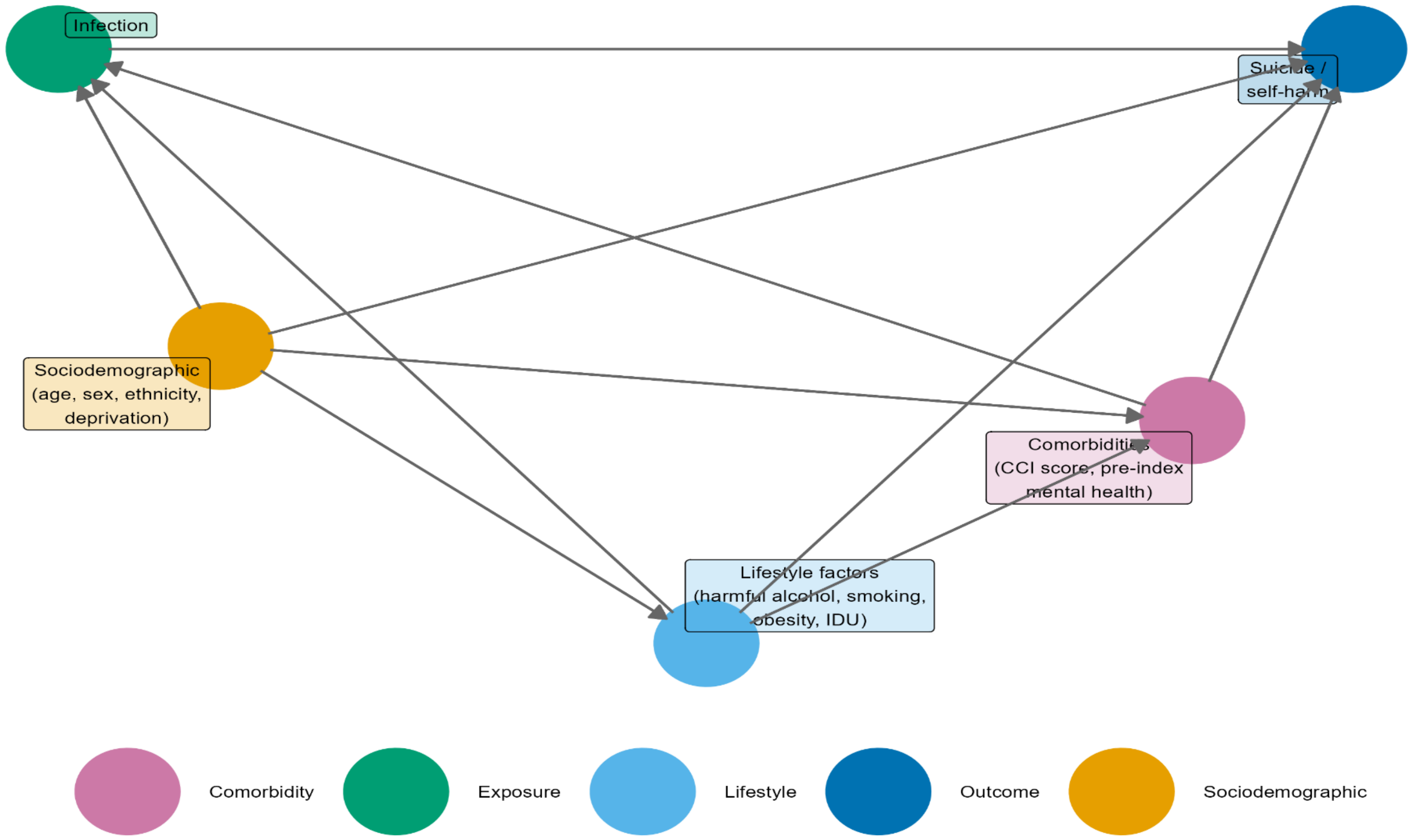


IDU = injecting drug use. IDU is included as a lifestyle covariate in the SSTI (skin and soft tissue infection) cohort only, reflecting its specific relevance as a risk factor for this infection type; it is not included in other cohorts.

CCI = Charlson Comorbidity Index, a weighted score of comorbid conditions. Pre-index mental health includes prior common mental disorders (CMD) and severe mental illness (SMI) recorded before the index date.

Sociodemographic factors (age, sex, practice, calendar period) are implicitly adjusted for via matched-set stratification; ethnicity and deprivation (IMD) are additionally adjusted for in the regression models.

**Supplementary Figure 3a. Flowchart for the data extraction process of the complete study cohorts**

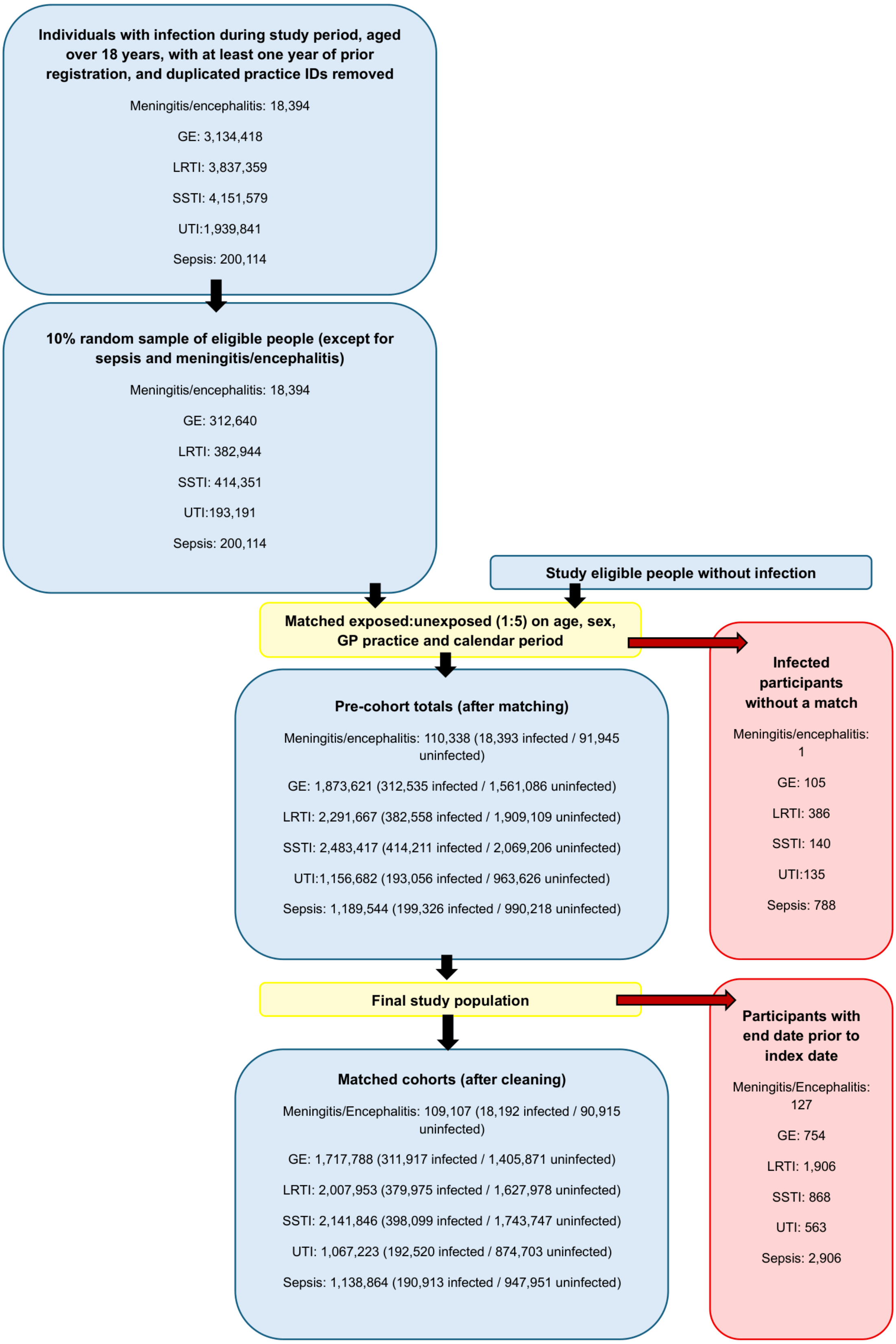

**Supplementary Figure 3b. Flowchart for the data extraction process of the analysis cohorts**

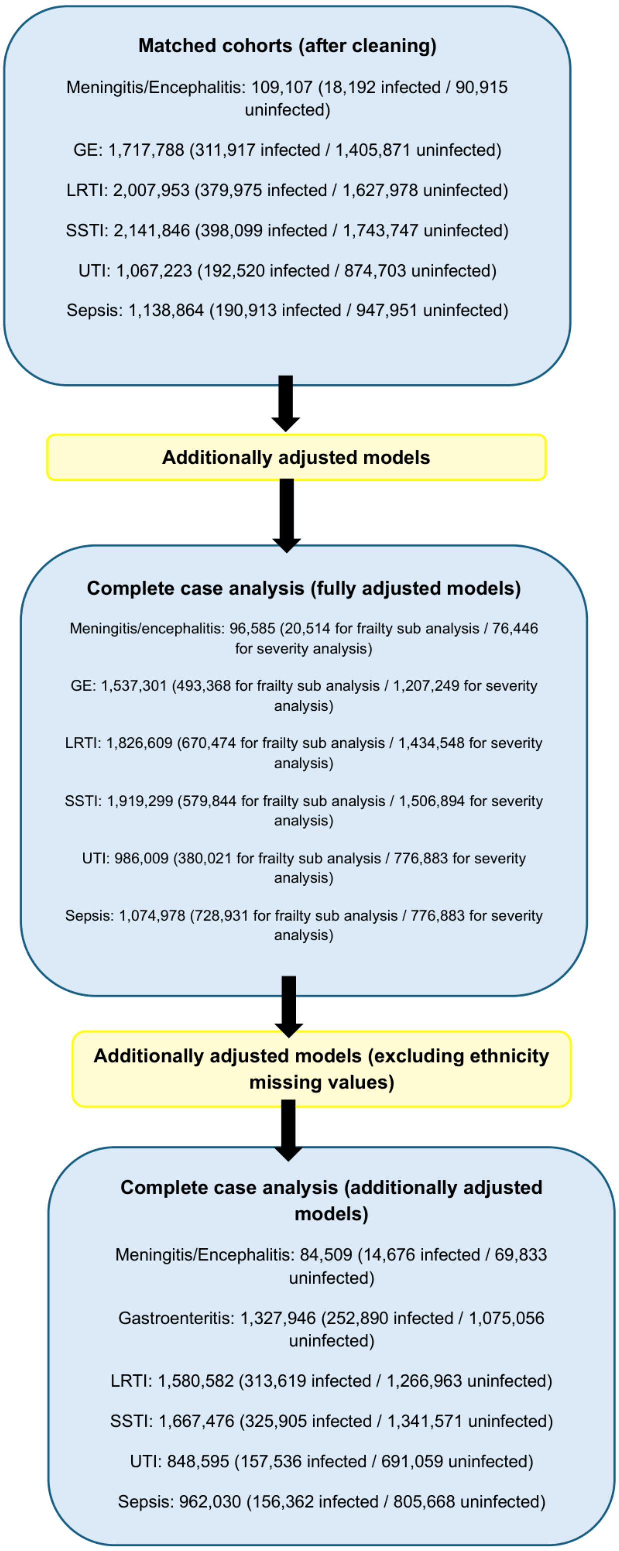

**Supplementary Figure 4. Suicidal outcome events by infection group**

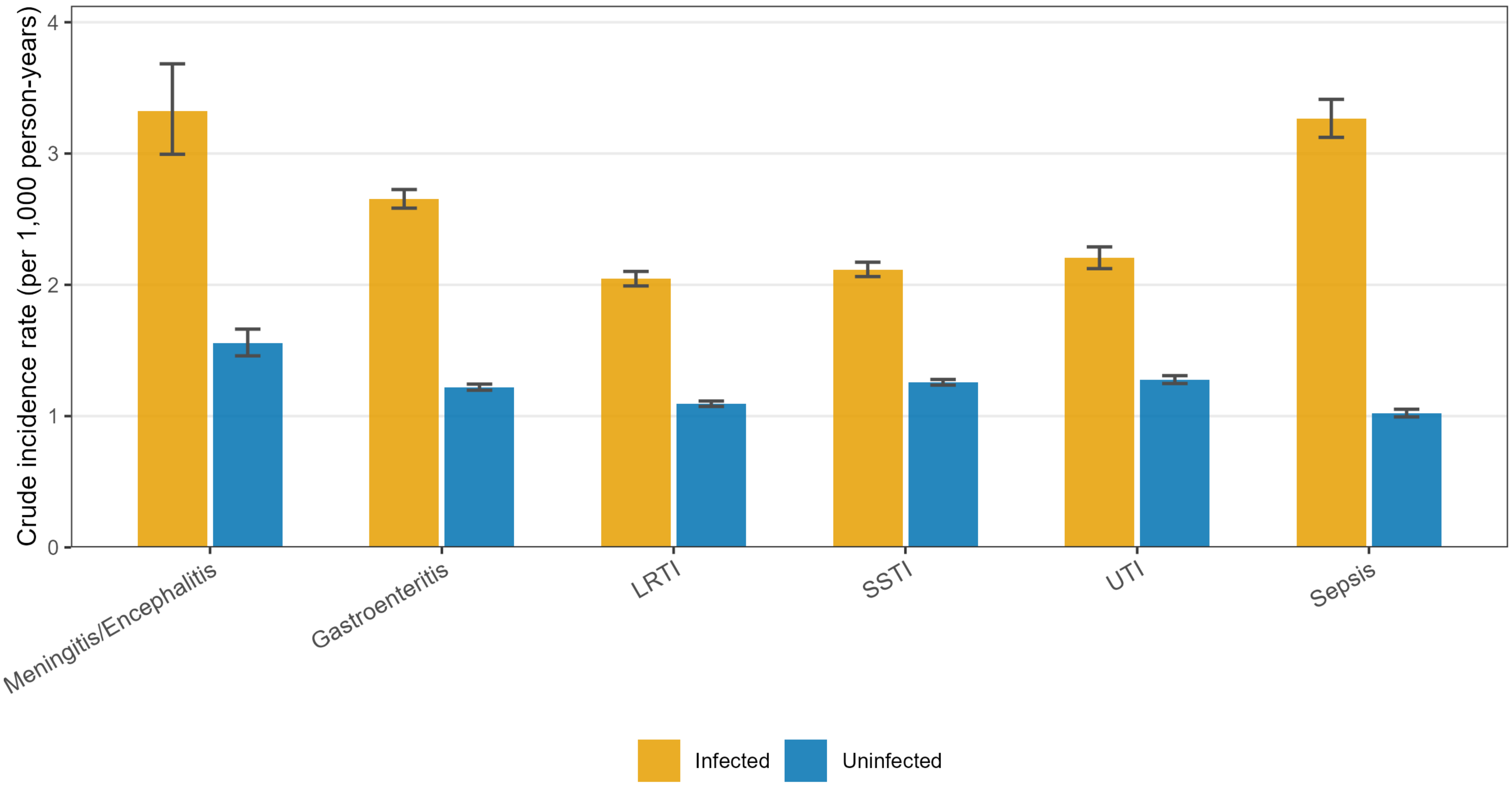


Rates = events / person-years × 1,000. Error bars = 95% CI (exact Poisson method).
Person-years calculated from index date to first outcome event or censoring.
Uninfected = matched comparators; rates account for person-time at risk in each group.

**Supplementary Figure 5. Kaplan-Meier plots comparing the survival for suicide/self-harm of individuals with and without an infection**

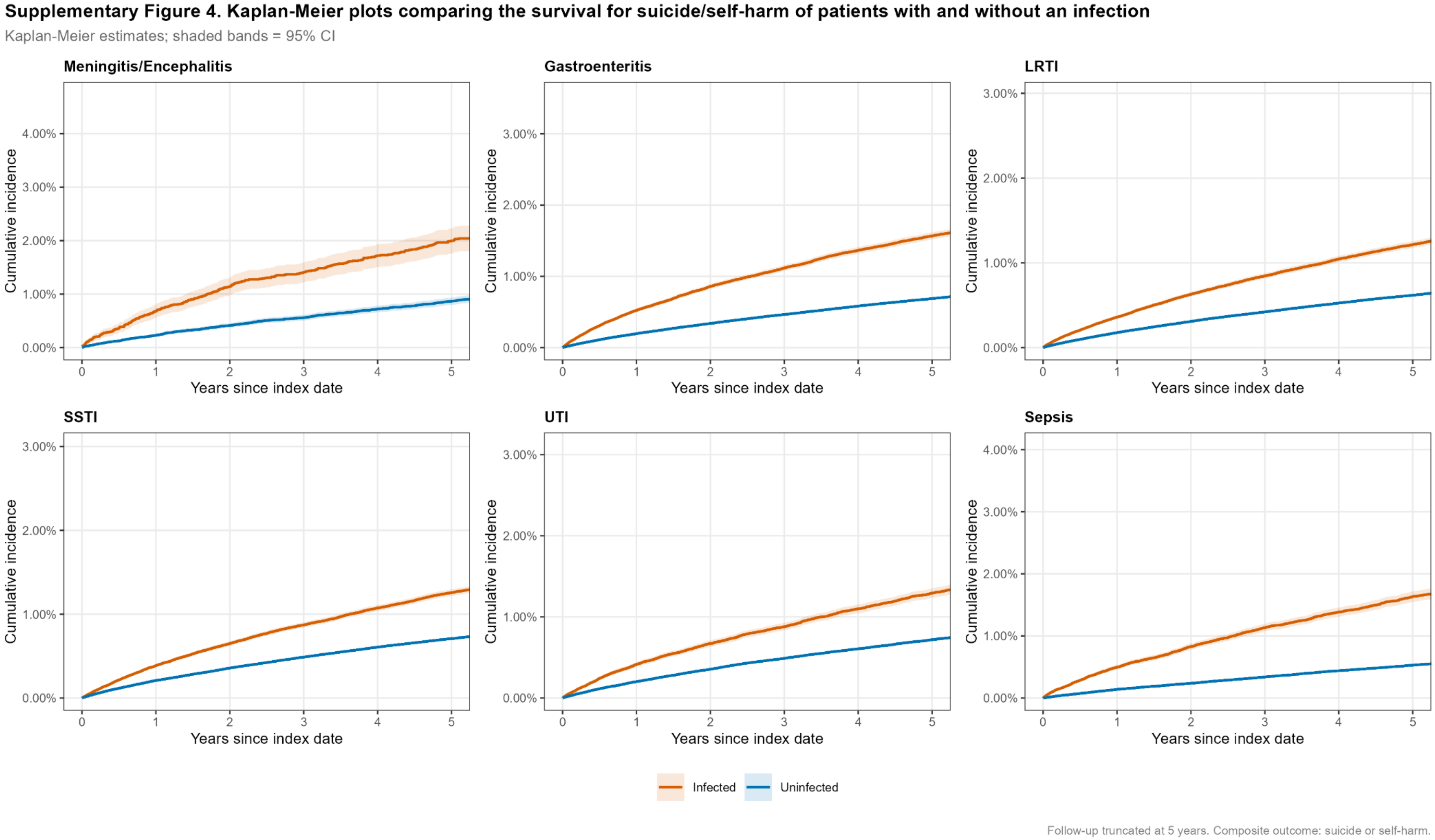

### Supplementary Figure 6. Model attenuation: hazard ratios for infection on suicide/self-harm across adjustment models

**Supplementary Figure 5. Model attenuation: hazard ratios for infection on suicide/self-harm across adjustment models**

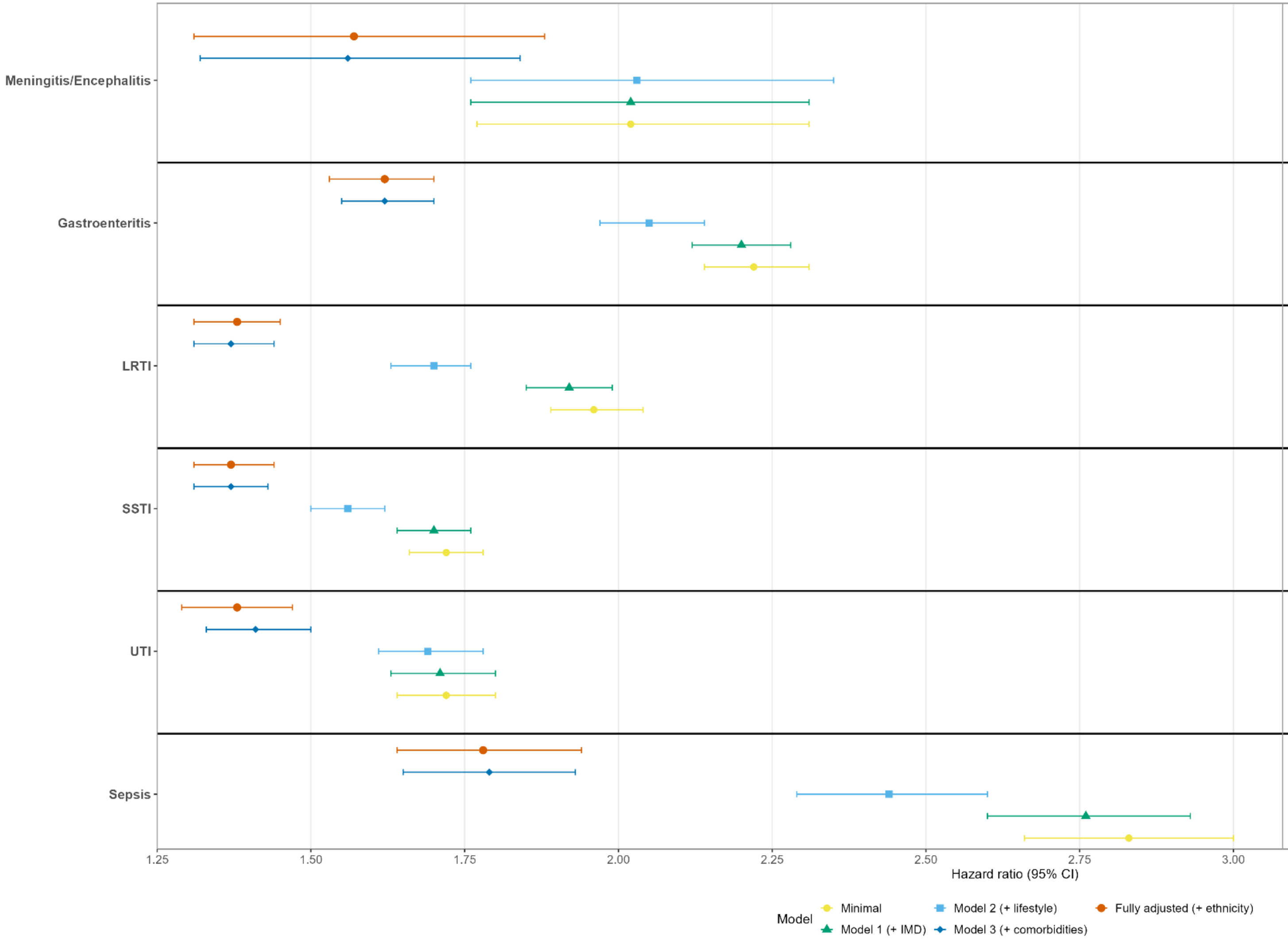


| | HR (95% CI) | n | Rate (per 1000 PY) | Model-derived RD (per 1000 PY) |
|---|---|---|---|---|
| Meningitis/Encephalitis | 1.57 (1.31–1.88) | 84,509 | | |
| | 1.56 (1.32–1.84) | 96,585 | | |
| | 2.03 (1.76–2.35) | 96,585 | 3.40 | 1.22 |
| | 2.02 (1.76–2.31) | 96,585 | | |
| | 2.02 (1.77–2.31) | 96,585 | | |
| Gastroenteritis | 1.62 (1.53–1.70) | 1,327,946 | | |
| | 1.62 (1.55–1.70) | 1,537,301 | | |
| | 2.05 (1.97–2.14) | 1,537,301 | 2.64 | 1.01 |
| | 2.20 (2.12–2.28) | 1,537,301 | | |
| | 2.22 (2.14–2.31) | 1,537,301 | | |
| LRTI | 1.38 (1.31–1.45) | 1,580,582 | | |
| | 1.37 (1.31–1.44) | 1,826,609 | | |
| | 1.70 (1.63–1.76) | 1,826,609 | 2.02 | 0.54 |
| | 1.92 (1.85–1.99) | 1,826,609 | | |
| | 1.96 (1.89–2.04) | 1,826,609 | | |
| SSTI | 1.37 (1.31–1.44) | 1,667,476 | | |
| | 1.37 (1.31–1.43) | 1,919,299 | | |
| | 1.56 (1.50–1.62) | 1,919,299 | 2.09 | 0.56 |
| | 1.70 (1.64–1.76) | 1,919,299 | | |
| | 1.72 (1.66–1.78) | 1,919,299 | | |
| UTI | 1.38 (1.29–1.47) | 848,595 | | |
| | 1.41 (1.33–1.50) | 986,009 | | |
| | 1.69 (1.61–1.78) | 986,009 | 2.19 | 0.64 |
| | 1.71 (1.63–1.80) | 986,009 | | |
| | 1.72 (1.64–1.80) | 986,009 | | |
| Sepsis | 1.78 (1.64–1.94) | 962,030 | | |
| | 1.79 (1.65–1.93) | 1,074,978 | | |
| | 2.44 (2.29–2.60) | 1,074,978 | 3.26 | 1.43 |
| | 2.76 (2.60–2.93) | 1,074,978 | | |
| | 2.83 (2.66–3.00) | 1,074,978 | | |

Points = HR; bars = 95% CI; dashed line = null (HR = 1). n = total participants in the analytic sample for each model (varies by model: minimal/Model 1-3 use the complete-case sample; fully adjusted uses the complete-ethnicity subset). Rate = observed crude incidence rate per 1,000 person-years (infected group, Model 3 CC sample). Model-derived RD = observed infected rate minus counterfactual uninfected rate (observed rate x 1/HR), per 1,000 person-years (Model 3).
